# To weight or not to weight? Studying the effect of selection bias in three large EHR-linked biobanks

**DOI:** 10.1101/2024.02.12.24302710

**Authors:** Maxwell Salvatore, Ritoban Kundu, Xu Shi, Christopher R Friese, Seunggeun Lee, Lars G Fritsche, Alison M Mondul, David Hanauer, Celeste Leigh Pearce, Bhramar Mukherjee

## Abstract

**Objective:** To explore the role of selection bias adjustment by weighting electronic health record (EHR)-linked biobank data for commonly performed analyses.

**Materials and methods:** We mapped diagnosis (ICD code) data to standardized phecodes from three EHR-linked biobanks with varying recruitment strategies: All of Us (AOU; n=244,071), Michigan Genomics Initiative (MGI; n=81,243), and UK Biobank (UKB; n=401,167). Using 2019 National Health Interview Survey data, we constructed selection weights for AOU and MGI to be more representative of the US adult population. We used weights previously developed for UKB to represent the UKB-eligible population. We conducted four common descriptive and analytic tasks comparing unweighted and weighted results.

**Results:** For AOU and MGI, estimated phecode prevalences decreased after weighting (weighted-unweighted median phecode prevalence ratio [MPR]: 0.82 and 0.61), while UKB’s estimates increased (MPR: 1.06). Weighting minimally impacted latent phenome dimensionality estimation. Comparing weighted versus unweighted PheWAS for colorectal cancer, the strongest associations remained unaltered and there was large overlap in significant hits. Weighting affected the estimated log-odds ratio for sex and colorectal cancer to align more closely with national registry-based estimates.

**Discussion:** Weighting had limited impact on dimensionality estimation and large-scale hypothesis testing but impacted prevalence and association estimation more. Results from untargeted association analyses should be followed by weighted analysis when effect size estimation is of interest for specific signals.

**Conclusion:** EHR-linked biobanks should report recruitment and selection mechanisms and provide selection weights with defined target populations. Researchers should consider their intended estimands, specify source and target populations, and weight EHR-linked biobank analyses accordingly.

## BACKGROUND AND SIGNIFICANCE

Electronic health record (EHR)-linked biobanks are repositories with biospecimen and/or related data that are linked to EHR and other forms of auxiliary data (e.g., medical and pharmacy claims, residential-level neighborhood characteristics).^1–4^ Many EHR-linked biobanks are non-probability samples (i.e., unknown selection probabilities^1,5–9^) drawn from a poorly defined *source population* (i.e., the population from which individuals are sampled). Because of their large sample size, linked multimodal data, immediacy and accessibility,^10–12^ researchers have been using EHR data *en masse* for scientific research (from 3,212 PubMed citations in 2013 to 9,824 in 2023). EHR-linked biobanks are increasingly prevalent and efforts like the Global Biobank Meta-analysis Initiative (GBMI)^13^ are facilitating collaboration across the globe.^14–18^

As the research community gets excited about amassing data, two fundamental questions must be asked: (a) who is in the study? and (b) what is the target population of interest? If biobanks are not representative of the target population, they are vulnerable to selection bias,^3,19–22^ a naïve analysis is not expected to align with the population truth.^23–25^ Handling selection bias is particularly challenging because the magnitude and direction of its impact on estimates are hard to determine,^26^ its effect cannot be mitigated by increasing sample size,^27,28^ and it can be coupled with other data imperfections.^3,29–32^ Moreover, contrary to previous arguments,^33,34^ recent evidence suggests that even genetic association analyses with inherited germline susceptibility factors can also be prone to selection bias.^35–38^

There are three common analytic approaches for handling selection bias: stratification,^24,25^ quantitative bias analysis,^24,39^ and, by far the most common, inverse probability (IP)-weighting.^3,4,21,25,29,35,40–43^ IP-weighting involves reweighting individuals in a given sample by the inverse of the estimated probability of their inclusion in the sample (relative to the target population) constructed as a function of variables that impact selection.^3,4,43^ The estimation of IP-weights relies on (a) access to representative individual-level data from the target population and (b) correct specification of the selection probability model. Representative data can be in the form of probability samples drawn from the target population like the National Health Interview Survey (NHIS; USA).^44^

Theory for and application of IP weights to EHR-linked biobank cohorts exists. Beesley and Mukherjee developed a framework for handling misclassification and selection bias using weighting procedures and demonstrated how individual-level data from external probabilistic samples can be used to estimate weights.^3,4^ Recently, van Alten and colleagues estimated lasso-based IP weights using United Kingdom Census Microdata to reweight the UK Biobank (UKB) sample to be more representative of the UKB-eligible population.^40^ Poststratification (PS)-weighting is an alternative that relies on summary-level data from the target population instead of individual level data.^45^

In this paper we consider three EHR-linked biobanks that have three different recruitment strategies/selection mechanisms: the National Institutes of Health All of Us Research Program (AOU),^7,46^ our University of Michigan’s Michigan Genomics Initiative (MGI),^8,47^ and the UKB.^48,49^ We explore the impact of the use of a set of selection weights on common descriptive (prevalence estimation, principal components analysis) and inferential (agnostic large-scale association testing, estimation of targeted association parameters) tasks in EHR data (**Figure S1**). First, we estimate selection weights in both US-based cohorts using NHIS data. Second, we characterize demographic and diagnostic (prevalences, latent dimensionality, partial correlation) data in AOU, MGI, and UKB, with and without selection weights. Third, we investigate how the use of weights impacts discovery in large-scale untargeted hypothesis testing by performing a phenome-wide association study (PheWAS). Fourth, we characterize the influence of weights on a targeted effect estimate in a fitted logistic regression model, using colorectal cancer as a sample phenotype. Finally, we discuss recommendations regarding the use of selection weights for practitioners conducting analyses in and across biobanks.

## MATERIALS AND METHODS

### Cohorts

#### AOU: All of Us

AOU started in 2018 with a goal of enrolling over one million adults via a combination of open invitations and a network of healthcare provider-based recruitment sites. Engagement efforts have focused on oversampling people from communities historically underrepresented in biomedical research based on 10 factors: age, sex, race/ethnicity, gender identity, sexual orientation, disability status, healthcare access, income, educational attainment, and geographic location.^7^ We considered these selection factors (except gender identity (not collected in NHIS) and disability status (significant missingness (∼61%) in AOU)) in the estimation of IP- and PS-based selection weights. As of January 1, 2024, there were over 760,000 participants, providing access to over 539,000 biosamples and 420,000 EHRs. The AOU subset used in these analyses consists of 244,071 participants with sociodemographic and ICD-9-CM/ICD-10-CM data as part of the curated data repository version 7 (Controlled Tier C2022Q4R9).

#### MGI: Michigan Genomics Initiative

The Michigan Medicine-based MGI (University of Michigan) began in 2012 recruiting adults primarily through appointments for procedures requiring anesthesia.^8^ It evolved to include sub-cohorts through metabolism, endocrinology and diabetes (MEND) and mental health (MHB) clinics and a wearables cohort enriched with hypertensive individuals (MIPACT). Age, sex, and race/ethnicity were considered selection factors. Additionally, cancer, diabetes and body mass index (BMI), anxiety and depression, and hypertension were selection mechanisms into the original cohort and these sub-cohorts, respectively, and were also used in selection weight estimation. As of September 2023, there were ∼100,000 consented participants in MGI with ongoing recruitment yielding ∼10,000 enrollments per year. The MGI subset used in these analyses consists of 81,243 participants (August 22, 2022, data pull) with demographic and ICD-9-CM/ICD-10-CM data.

#### UKB: UK Biobank

The UKB recruited more than 500,000 adults aged 40-69 by mailing over 9 million invitations to homes within ∼40 kilometers of 22 assessment centers across the UK. Following evidence of healthy volunteer bias,^9^ van Alten and colleagues developed a set of generic weights to reweight the UKB sample to the UKB-eligible population using UK Census Microdata.^40^ Using an array of sociodemographic characteristics – age, sex, race/ethnicity, educational attainment, employment status, location of residence, tenure of dwelling, number of cars in household, self-reported health, and one-person household status – they estimate lasso regression-based IP-weights.^40^ These weights were used in this paper. The UKB subset used in these analyses consists of 401,167 participants with sociodemographic and ICD-10 code data remaining after phenome curation (**Figure S2**).

### Phenome curation

For all cohorts, ICD-9-CM and ICD-10(-CM) codes were recoded into up to 3,612 phecodes across 18 phecode categories (i.e., phecodes, or “PheWAS codes”^50^), using the phecode X mapping tables (downloaded from GitHub^51^ on 6 September 2023) and the PheWAS R package (version 0.99.6-1).^52^ Cases were defined as individuals with a single occurrence of a corresponding phecode. There were 3,493, 3,354, and 2,660 phecodes defined in AOU, MGI, and UKB, respectively; we restricted our analyses to the 2,042 phecodes that had at least 20 cases in all three cohorts. Flowcharts depicting sample size changes following filtering and ICD-to-phecode mapping for all cohorts are shown in **Figure S2**. Phecode-derived trait mappings are shown in **Table S1**.

### Weight estimation

#### Inverse probability weighting

We constructed IP-weights, which require individual-level data in the target population, in the US-based cohorts. To do this, we used the 2019 NHIS, a probabilistic sample of US adults with self-reported health information. We estimated selection probabilities, *ψ*, using a simplex regression framework based on the Beta regression approach to weight estimation described in Kundu and colleagues^21^:

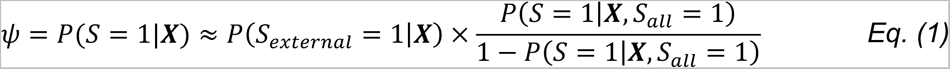

where, assuming there is no overlap between the internal and external data, *S* is an inclusion indicator in the internal cohort (i.e., AOU or MGI), *S_external_* is an indicator for inclusion in the external cohort (i.e., NHIS), *S_all_* is an indicator for inclusion in either cohort, and ***X*** are selection factors as listed in the **Cohorts** section (**Figure 1**). We estimated the first term, P(*S_external_* = 1|***X)***, by fitting a simplex regression model for the known design probabilities using NHIS data. We estimated the numerator of the second term, *P*(*S* =1|***X***, *S_all_* = 1), using a logistic regression model using both internal and external data.

**Figure 1.**
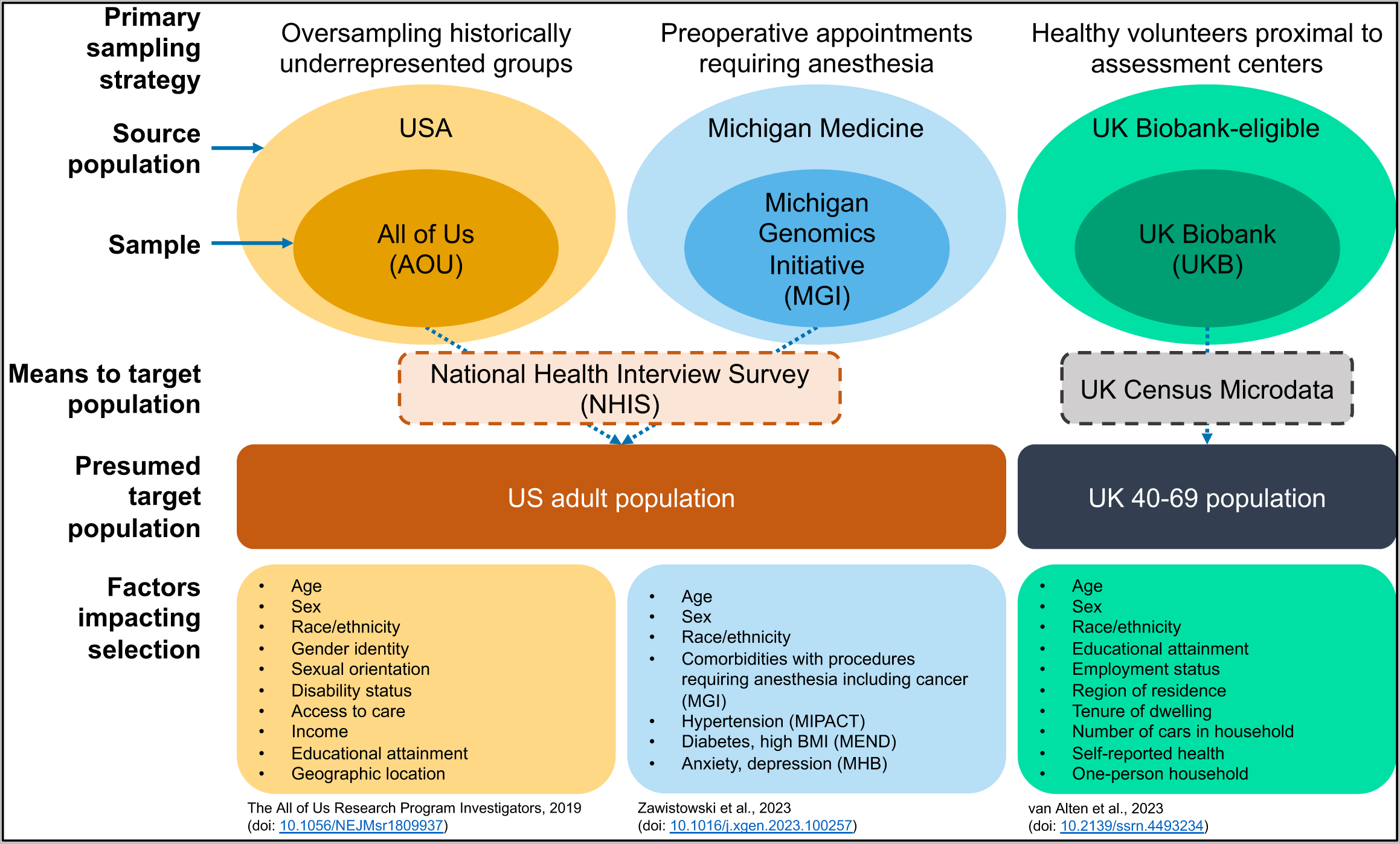
Schematic representation of the All of Us, the Michigan Genomics Initiative, and the UK Biobank cohorts, their sampling strategies, potential target populations, and selection factors. All three cohorts are non-probability samples of their source populations for different reasons: oversampling, procedures requiring anesthesia, and healthy volunteers, respectively. External data like NHIS or UK Census Microdata can be used in selection weight construction to make inferences regarding presumed target populations. Factors known to influence recruitment strategy or eligibility criteria are listed.

In AOU, we flexibly selected ***X*** by splitting the data in half and fitting a lasso-penalized logistic regression model on ***X*** and all possible pairwise interactions using the glmnet R package (version 4.1-8). Using 10-fold cross-validation, we selected λ such that the error is within 1 standard error of the minimum to result in a parsimonious model. The selected terms were then used as the final set of ***X*** to estimate IP weights in the other half of the data as described above.

#### Poststratification

Using weighted NHIS data, the poststratification (PS)-weights were calculated using:

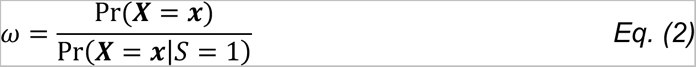

where ***X*** are the set of selection variables, and ***S*** is an indicator for membership in the internal sample (i.e., AOU or MGI). IP- and PS-weights were winsorized at the 2.5^th^ and 97.5^th^ percentile. Variable definitions are described in **Table S2,** and additional details of IP- and PS-weight estimation are described in **Supplementary Methods**.

A schematic representing the cohorts and their source populations, sampling strategies, presumed target populations, external data for weighting, and selection factors is presented in **Figure 1**.

### Statistical analyses

First, we obtained crude unweighted and IP-weighted estimates of *prevalences*. These are calculated as the number of cases over the number of individuals in the respective biobanks. For sex-specific phecodes, only individuals with the corresponding sex are considered.

Second, we estimated the latent *dimensionality* of the phenome by conducting unweighted and IP-weighted principal components analyses (PCA). We used the number of principal components explaining 95% and 99% of the cumulative variation in the data to represent its dimensionality. Additionally, we explored partial *correlations*, described in **Supplementary Methods**.

Third, we conducted a colorectal cancer (phecode CA_101.41) PheWAS to illustrate large-scale *hypothesis testing*. Here, the interest was in obtaining the test statistic and corresponding *p-value*. PheWAS were adjusted for age, sex, and length of EHR follow-up.

Fourth, we estimated the association between biological sex and colorectal cancer, where the interest was in estimating the log-odds ratio. The female-colorectal cancer association was selected because it is known to be negative (recent log-odds ratio estimate approximations range from -0.414 to -0.271) in the US^53^ and the UK.^54^ For hypothesis testing and targeted association analyses, after performing a weighted or unweighted analysis within each cohort, we conducted a meta-analysis across three cohorts by using inverse variance weights and a fixed effect model using the meta R package (version 6.5-0) (**Figure S3**).^55^ Additional data preparation detail is described in **Supplementary Methods**.

### Software

All data cleaning, manipulation, and analysis were conducted using R version 4.2.2. Code is publicly available: https://github.com/maxsal/biobank_selection_weights.

## RESULTS

### Descriptive characteristics

Of 244,071 AOU participants, 62.2% were female, with a mean (standard deviation (SD)) age of 54.0 (17.3) years old (**Table 1**). Additionally, 55.4% were non-Hispanic White, and 27.1% had a qualifying cancer phecode in their EHR. Of 81,243 MGI participants, 53.8% were female with a mean age of 56.3 (17.0) years old. Most of MGI was non-Hispanic White (83.1%) and 49.2% had a cancer diagnosis on their EHR. MGI had substantially more EHR data points per person than AOU as measured by encounters per person (mean 103 in MGI vs. 32 in AOU), unique phecodes per person (77 vs. 72), and years of follow-up per person (9.9 vs. 9.3). Both IP- and PS-weighting brought AOU and MGI closer to NHIS-based estimates of the US population with respect to age (47.7 years old), sex (51.7% female), and race/ethnicity (63.2% non-Hispanic White).

**Table 1.** Descriptive characteristics of the Michigan Genomics Initiative, the UK Biobank, and All of Us. For unweighted metrics, mean (standard deviation) and percent (n) are provided for continuous and categorical/binary variables, respectively. For weighted metrics, mean (standard error) and percent (standard error) are provided for continuous and categorical/binary variables, respectively.

|  | NHIS<sup>a</sup> (weighted) | All of Us (AOU)<sup>b</sup> |  |  | Michigan Genomics Initiative (MGI) |  |  | UK Biobank (UKB)<sup>c</sup> |  |  |
| --- | --- | --- | --- | --- | --- | --- | --- | --- | --- | --- |
|  |  | Unweighted | IP-weighted | PS-weighted | Unweighted | IP-Weighted | PS-weighted | Eligible population | Unweighted | IP-weighted |
| N |  | 244,071 | - | - | 81,243 | - | - | - | 401,167 | - |
| *Demographics* |  |  |  |  |  |  |  |  |  |  |
| Age (continuous)<sup>d</sup> | 47.7 (0.2) | 54.01 (17.27) | 49.8 (0.1) | 51.1 (0.1) | 56.34 (16.99) | 45.5 (0.1) | 45.6 (0.1) | 54.78 (0.01) | 57.67 (8.02) | 54.9 (0.0) |
| Age<sup>d</sup> |  |  |  |  |  |  |  |  |  |  |
| 0-9 | 0.0 (0.0) | 0.0 (56) | 0.0 (0.0) | 0.0 (0.0) | 0.0 (1) | 0.0 (0.0) | 0.0 (0.0) | - | 0.0 (0) | 0.0 (0.0) |
| 10-19 | 3.4 (0.2) | 0.8 (2,007) | 1.1 (0.1) | 0.8 (0.0) | 0.6 (472) | 2.2 (0.1) | 2.2 (0.1) | - | 0.0 (0) | 0.0 (0.0) |
| 20-29 | 17.6 (0.3) | 10.2 (24,907) | 13.0 (0.2) | 11.1 (0.1) | 8.7 (7,067) | 21.9 (0.3) | 21.6 (0.3) | - | 0.0 (0) | 0.0 (0.0) |
| 30-39 | 17.1 (0.3) | 14.2 (34,719) | 19.7 (0.2) | 19.1 (0.1) | 10.6 (8,636) | 18.4 (0.3) | 18.7 (0.3) | - | 0.0 (3) | 0.0 (0.0) |
| 40-49 | 16.0 (0.3) | 14.0 (34,080) | 16.6 (0.2) | 20.0 (0.1) | 13.5 (10,928) | 19.7 (0.3) | 20.0 (0.3) | - | 21.0 (84,174) | 34.1 (0.1) |
| 50-59 | 16.4 (0.3) | 19.2 (46,937) | 18.4 (0.2) | 14.8 (0.1) | 19.9 (16,127) | 14.1 (0.2) | 13.9 (0.2) | - | 32.0 (128,374) | 31.5 (0.1) |
| 60-69 | 15.4 (0.2) | 21.4 (52,112) | 17.5 (0.2) | 17.1 (0.1) | 23.1 (18,801) | 13.1 (0.2) | 12.9 (0.2) | - | 46.3 (185,741) | 33.8 (0.1) |
| 70-79 | 9.4 (0.2) | 15.4 (37,532) | 10.8 (0.1) | 13.2 (0.1) | 17.4 (14,108) | 8.0 (0.1) | 8.0 (0.2) | - | 0.7 (2,874) | 0.6 (0.0) |
| 80+ | 4.9 (0.1) | 4.8 (11,721) | 2.8 (0.1) | 3.9 (0.1) | 6.3 (5,103) | 2.6 (0.1) | 2.8 (0.1) | - | 0.0 (0) | 0.0 (0.0) |
| Sex |  |  |  |  |  |  |  |  |  |  |
| Male | 48.3 (0.4) | 37.8 (92,315) | 45.5 (0.2) | 46.8 (0.2) | 46.2 (37,541) | 48.6 (0.3) | 49.9 (0.3) | 49.2 (0.1) | 44.7 (179,135) | 47.2 (0.1) |
| Female | 51.7 (0.4) | 62.2 (151,756) | 54.5 (0.2) | 53.2 (0.2) | 53.8 (43,702) | 51.4 (0.3) | 50.1 (0.3) | 50.8 (0.1) | 55.3 (222,032) | 52.8 (0.1) |
| Race/Ethnicity |  |  |  |  |  |  |  |  |  |  |
| Asian | 5.9 (0.3) | 2.7 (6,535) | 2.9 (0.1) | 3.0 (0.1) | 3.7 (2,983) | 11.4 (0.3) | 11.4 (0.3) | - | 2.2 (8,898) | 4.6 (0.1) |
| Black or African American | 11.8 (0.4) | 18.5 (45,243) | 14.6 (0.2) | 15.3 (0.1) | 6.1 (4,959) | 11.1 (0.2) | 11.6 (0.3) | - | 1.6 (6,385) | 2.8 (0.1) |
| Hispanic | 16.5 (0.6) | 18.9 (46,032) | 13.5 (0.1) | 13.4 (0.1) | 2.7 (2,179) | 6.4 (0.2) | 6.5 (0.2) | - | - | - |
| Other/Unknown | 2.6 (0.3) | 4.5 (11,096) | 3.2 (0.1) | 3.6 (0.1) | 4.4 (3,578) | 5.9 (0.2) | 6.1 (0.2) | - | 2.0 (8,094) | 1.7 (0.0) |
| White | 63.2 (0.8) | 55.4 (135,165) | 65.9 (0.2) | 64.7 (0.1) | 83.1 (67,544) | 65.1 (0.4) | 64.4 (0.4) | 87.0 (0.05) | 94.2 (377,790) | 90.9 (0.1) |
| BMI (continuous) | 27.8 (0.0) | 30.02 (7.67) | 29.9 (0.0) | 30.0 (0.0) | 29.94 (7.26) | 28.0 (0.1) | 28.1 (0.1) |  | 27.64 (4.90) | 27.9 (0.0) |
| BMI |  |  |  |  |  |  |  |  |  |  |
| Underweight (<18.5) | 1.6 (0.1) | 1.3 (2,977) | 1.3 (0.1) | 1.1 (0.0) | 1.1 (865) | 1.9 (0.1) | 1.7 (0.1) | - | 0.5 (2,052) | 0.6 (0.0) |
| Healthy [18.5, 25) | 31.4 (0.3) | 25.6 (59,653) | 26.7 (0.2) | 26.1 (0.1) | 24.7 (20,025) | 35.9 (0.3) | 34.8 (0.3) | - | 31.0 (123,485) | 30.1 (0.1) |
| Overweight [25, 30) | 33.0 (0.3) | 30.6 (71,174) | 30.5 (0.2) | 30.9 (0.1) | 32.0 (25,954) | 33.9 (0.3) | 33.8 (0.3) | - | 42.5 (169,172) | 41.4 (0.1) |
| Obese [30+) | 31.2 (0.4) | 42.5 (99,024) | 41.5 (0.2) | 41.9 (0.2) | 42.3 (34,285) | 28.4 (0.3) | 29.7 (0.3) | - | 26.0 (103,779) | 27.9 (0.1) |
| Anxiety | 6.1 | 32.0 (78,018) | 30.5 | 23.7 | 35.3 (28,690) | 8.5 | 8.0 | - | 5.5 (22,047) | 6.3 |
| Cancer<sup>e</sup> | 9.5 | 27.1 (66,124) | 22.9 | 24.5 | 49.2 (39,984) | 8.3 | 9.8 | - | 25.9 (103,996) | 23.7 |
| Depression | 7.0 | 25.7 (62,689) | 23.8 | 19.2 | 31.6 (25,634) | 7.5 | 7.3 | - | 0.2 (712) | 0.2 |
| Diabetes | 8.0 | 23.0 (56,210) | 18.7 | 42.6 | 23.1 (18,790) | 8.0 | 7.4 | - | 10.1 (40,353) | 11.4 |
| Hypertension | 31.7 | 47.6 (116,156) | 40.8 | 30.4 | 50.7 (41,182) | 29.9 | 29.9 | - | 33.7 (135,113) | 32.1 |
| *EHR characteristics* |  |  |  |  |  |  |  |  |  |  |
| Encounters per person (unique)<sup>f</sup> |  |  |  |  |  |  |  |  |  |  |
| Minimum | - | 1 | - | - | 1 | - | - | - | 1 | - |
| Median | - | 19 | - | - | 54 | - | - | - | 3 | - |
| Mean | - | 32 | - | - | 103 | - | - | - | 4 | - |
| Maximum | - | 487 | - | - | 1833 | - | - | - | 92 | - |
| Unique phecodes per person<sup>f</sup> |  |  |  |  |  |  |  |  |  |  |
| Minimum | - | 1 | - | - | 1 | - | - | - | 1 | - |
| Median | - | 52 | - | - | 55 | - | - | - | 12 | - |
| Mean | - | 72 | - | - | 77 | - | - | - | 17 | - |
| Maximum | - | 573 | - | - | 591 | - | - | - | 212 | - |
| Length of follow-up (years)<sup>f</sup> |  |  |  |  |  |  |  |  |  |  |
| Minimum | - | 0.0 | - | - | 0.0 | - | - | - | 0.0 | - |
| Median | - | 7.4 | - | - | 8.1 | - | - | - | 7.0 | - |
| Mean | - | 9.3 | - | - | 9.9 | - | - | - | 7.5 | - |
| Maximum | - | 41.5 | - | - | 43.5 | - | - | - | 22.0 | - |

Abbreviations: -, data unavailable in cohort; AOU, All of Us; IP, inverse probability; MGI, Michigan Genomics Initiative; NHIS, National Health Interview Survey; PS, poststratification; UKB, UK Biobank
<sup>a</sup> 2019 NHIS data
<sup>b</sup> All of Us data restricted to those who are present in the phenome.
<sup>c</sup> The UKB weighting procedure and definitions of the UKB-eligible population are described in van Alten and colleagues (doi: 10.1101/2022.05.16.22275048). Further, estimates presented here of the UKB-eligible population are taken from van Alten and colleagues.
<sup>d</sup> Age defined as age at last EHR encounter in AOU and MGI, age at consent in UKB
<sup>e</sup> Presence of any qualifying cancer phecode in phenome
<sup>f</sup> UK Biobank data only provided first occurrence of each unique phecode. This means that individuals could have encounters that were not reported or extend beyond their last encounter if no unique diagnoses were recorded at that visit.
Notes: Descriptions of how cancer, diabetes, coronary heart disease, and depression can be found in **Tables S1** and **S2**.

Of the 401,167 participants in UKB, 55.3% were female and their mean age was 57.7 (8.0) years old. Additionally, they were 94.2% White, and 25.9% had a qualifying cancer phecode on their EHR. The application of the IP weights resulted in a cohort that was reflective of the UKB-eligible population with respect to age (54.9 weighted vs 54.8 UKB-eligible), sex (50.8% female weighted vs 50.8% female UKB-eligible), and race/ethnicity (90.9% White weighted vs 87.0% White UKB-eligible).

### Phecode prevalences

#### Within cohort comparison

In AOU, unweighted phecode prevalences ranged from <0.01% to 52.07% with a median of 0.40%, while weighted prevalences ranged from 0% to 46.86% with a median of 0.20%. Weighted-to-unweighted phecode prevalence ratios (PR; **Figure 2A**) were downweighted (i.e., below 1) phenome-wide with a median PR (MPR) is 0.82. In MGI, unweighted prevalences ranged from <0.01% to 50.69% with a median of 0.33%, while weighted prevalences ranged from 0% to 43.12% with a median of 0.21%. Weighting tended to downweight prevalences with an MPR of 0.61 (**Figure 2B**). In UKB, unweighted prevalences spanned <0.01% to 33.68%, with a median of 0.06%, while weighted prevalences spread from 0% to 32.12% with a median of 0.07%. Weighting tended to upweight prevalences with a MPR of 1.06 (**Figure 2C**).

**Figure 2.**
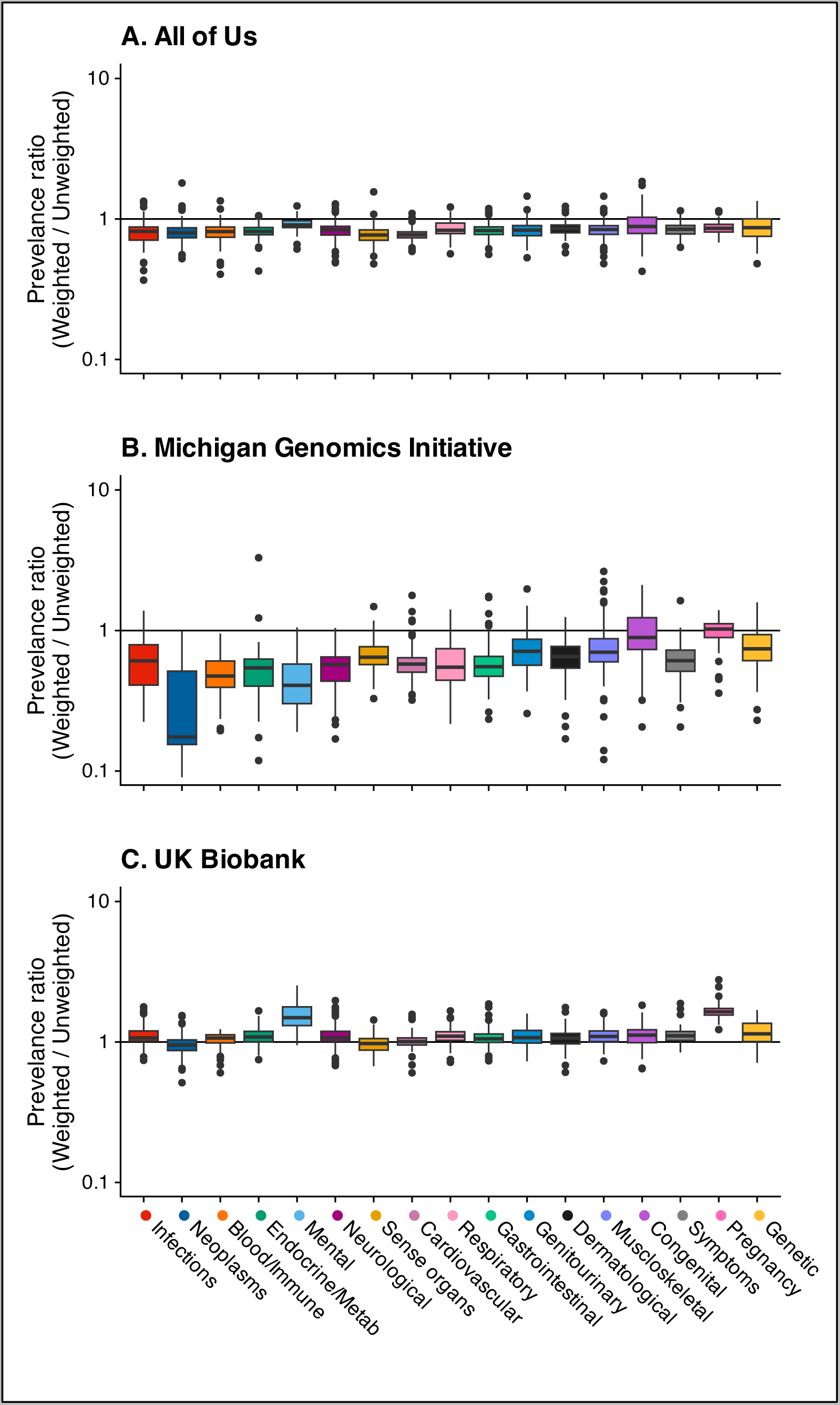
Side-by-side boxplots of the inverse probability (IP)-weighted over unweighted phecode prevalence ratios **within cohorts** by 17 defined phecode categories. Panel A shows the ratio of IP-weighted/unweighted prevalences in AOU, panel B shows the ratio of IP-weight/unweighted prevalences in MGI, and panel C shows the ratio of IP- weighted/unweighted prevalances in UKB. IP-weights were used in AOU and MGI and IP-weights described in van Alten et al.^40^ were used in UKB.

#### Across cohorts comparison

Comparing unweighted phecode prevalences, MGI over AOU (**Figure 3A**), we calculated a median and mean PR of 1.15 and 1.70, respectively. On average, 13 of 17 phecode categories had higher prevalences in MGI compared to AOU except for infections, dermatological, pregnancy, and mental categories (MPRs 0.97, 0.92, 0.88, and 0.74, respectively). Neoplasms were substantially more common in MGI (MPR 2.69). After IP-weighting both cohorts (**Figure 3D**), median and mean PRs were 0.81 and 1.23, respectively. Only congenital and genetic (MPRs 1.70, 1.02, respectively) phecodes remained more common in MGI after weighting.

**Figure 3.**
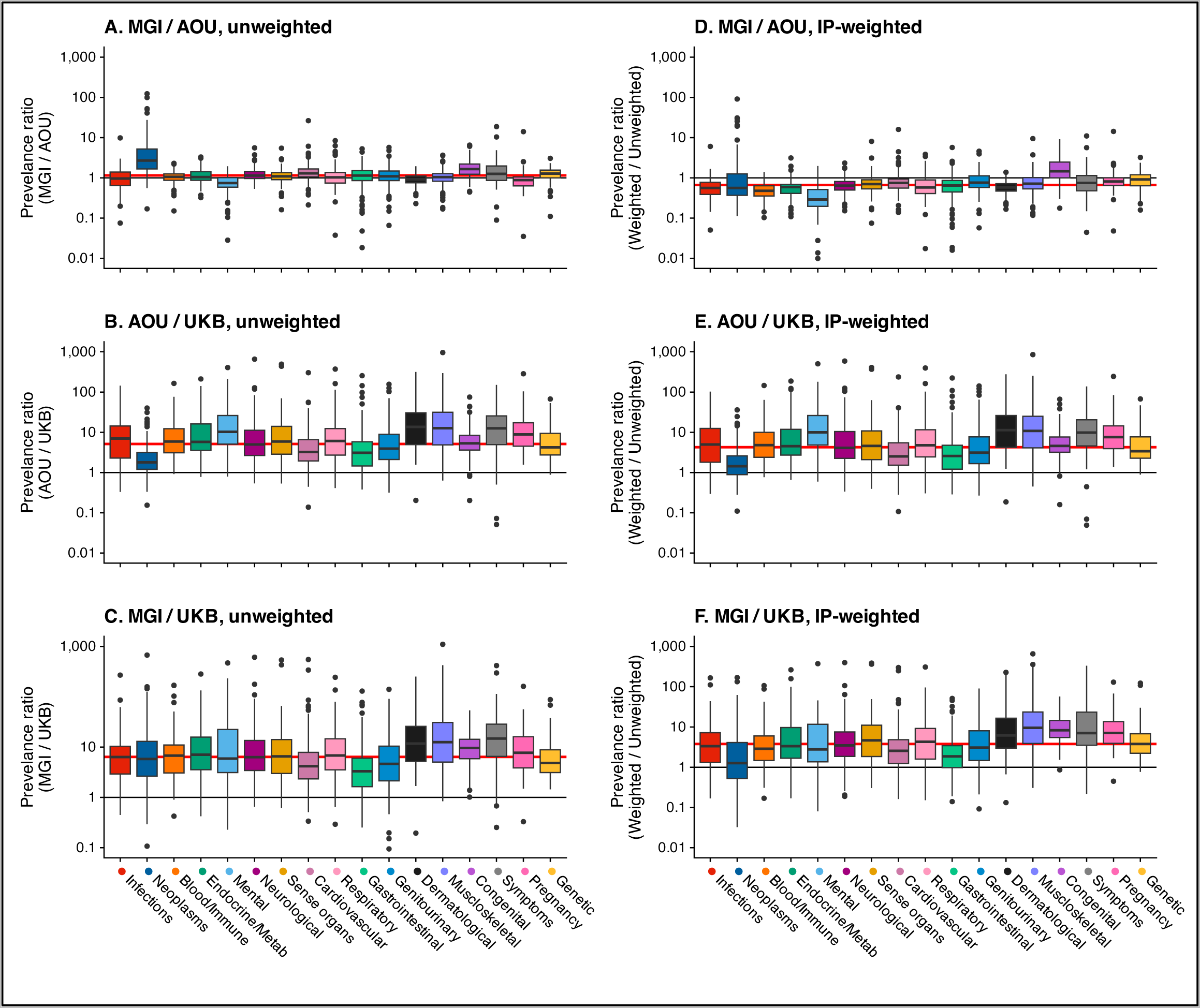
Side-by-side boxplots of the unweighted and inverse probability (IP)-weighted phecode prevalence ratios **across cohorts** by 17 defined phecode categories. Panel A shows the ratio of unweighted prevalences in MGI over AOU, panel B shows the ratio of unweighted prevalences in AOU / UKB, and panel C shows the ratio of unweighted prevalances in MGI / UKB. Panel D shows the ratio of IP-weighted prevalences in MGI over AOU, panel E shows the ratio of IP-weighted prevalences in AOU / UKB, and panel F shows the ratio of IP-weighted prevalances in MGI / UKB. The horizontal red line indicates the median phenome-wide prevalence ratio value. IP-weights were used in AOU and MGI and IP-weights described in van Alten et al.^40^ were used in UKB.

Using unweighted data (**Figure 3B** and **C**), phecodes in AOU and MGI were more common than in UKB (MPR: AOU/UKB 5.12; MGI/UKB: 6.37). After IP-weighting (**Figure 3E** and **F**), phecodes in AOU and MGI were still magnitudes more common than in UKB (M PR: AOU/UKB: 3.87; MGI/UKB 3.39).

### Phenome structure: PCA to estimate the effective number of phenotypes

The latent dimensionality of the diagnostic phenome (n = 2,042) was estimated using PCA in AOU, MGI, and UKB (**Table 2**; shown graphically in **Figure S4**). Within cohorts, weighting nominally decreased the number of PCs explaining 95% of cumulative variation (CV) in AOU and MGI (from 732 to 711 in AOU; from 752 to 729 in MGI) and nominally increased in UKB (from 553 to 569). This trend was the same at the 99% of CV threshold (from 1,262 to 1,236 in AOU; from 1,293 to 1,258 in MGI; from 1,065 to 1,080 in UKB). Dimensionality of the UKB data was noticeably smaller than the US-based cohorts with higher phecode prevalences (e.g., at the 95% CV threshold, 569 PCs weighted UKB phenome vs. 711 and 729 PCs in AOU and MGI, respectively).

**Table 2.** Number of principal components by proportion of cumulative variation (CV) in diagnostic phenome (n = 2,042) explained by cohort.

|  | **95% CV explained** |  | **99% CV explained** |  |
| --- | --- | --- | --- | --- |
|  | **Unweighted** | **Weighted** | **Unweighted** | **Weighted** |
| **All of Us** | 732 | 711 | 1,262 | 1,236 |
| **Michigan Genomics Initiative** | 752 | 729 | 1,293 | 1,258 |
| **UK Biobank** | 553 | 569 | 1,065 | 1,080 |

Weighted results were conducted using inverse probability (IP)-weights. Out of 2,042 phecodes with at least 20 cases in all three cohorts.

As a supplemental exploration (**Section S1**), we calculated unweighted and weighted partial correlations. Partial correlations were visualized as network graphs for AOU, MGI, and UKB in **Figures S5**, **S6**, and **S7**, respectively, and did not show noticeable differences after weighting. Distributions of unweighted (**Figure S8**) and weighted (**Figure S9**) partial correlations showed that cohorts with higher phecode prevalences (e.g., MGI) had slightly stronger correlations than those with lower phecode prevalences (e.g., UKB).

### Large-scale hypothesis testing: an “untargeted” PheWAS for colorectal cancer

In AOU, there were 25 phenome-wide significant hits in the unweighted PheWAS across 6 categories (**Figure 4A**). After IP-weighting, there were only 5 hits, all neoplasms (**Figure 4D**) – the same top 5 hits as in the unweighted PheWAS. In MGI, there were 9 phenome-wide significant hits in the unweighted PheWAS across 2 categories (**Figure 4B**). After IP-weighting, there were 26 hits across 4 categories (**Figure 4E**). Of the 9 unweighted hits, only 3 were identified in the IP-weighted PheWAS. The IP-weighted PheWAS identified 23 hits that were not identified in the unweighted PheWAS. In UKB, there were 60 phenome-wide-significant hits in the unweighted PheWAS across 11 categories (**Figure 4C**). After IP-weighting, there were 34 hits across 8 categories (**Figure 4F**). Of the 60 unweighted hits, 30 were also identified in the weighted PheWAS. There were 4 new gastrointestinal hits in the weighted PheWAS. Overlaps in phenome-wide significant hits across weighting strategies within cohort are shown as Venn diagrams in **Figure S10A-C**.

**Figure 4.**
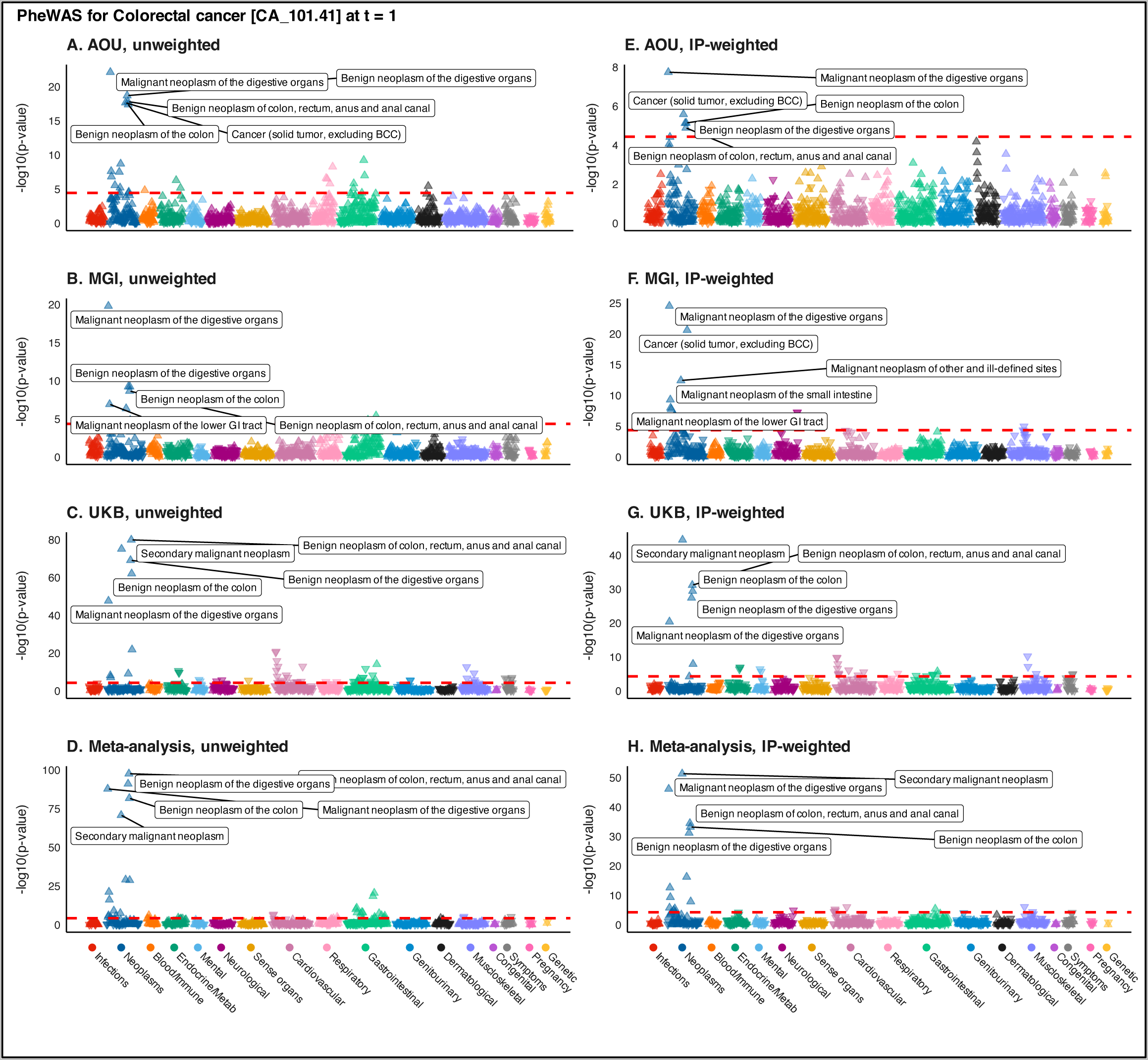
Manhattan plots summarizing unweighted (panels A-C) and inverse probability (IP)-weighted (panels E-G) phenome-wide association studies (PheWAS) for colorectal cancer in All of Us, the Michigan Genomics Initiative, and UK Biobank using 1:2 case:non-case matched data restricted to one year prior to initial diagnosis. Panels D and H show the unweighted and IP-weighted meta-analysis PheWAS, respectively. The dashed red line represents the Bonferroni-corrected p-value threshold (-log10(0.05/number of traits)). The five traits with the smallest p-values are labeled. The upward (downward) orientation of the triangle indicates a positive (negative) association. Plots corresponding to poststratification-weighted PheWAS are presented in supplementary **Figure S12**.

Of the 96 unique hits identified in any unweighted or IP-weighted PheWAS, 21.9% (n = 21) appeared only in IP-weighted PheWAS. Most of these hits found only in weighted PheWAS were neoplasms (11) with others belonging to the gastrointestinal (4), neurological (3), mental (1), and musculoskeletal (2) categories. The only hit identified in all three IP-weighted PheWAS (CA_101: Malignant neoplasm of the digestive organs) was also identified in all three unweighted PheWAS. Of the 21 hits only identified in IP-weighted PheWAS, 71.4% (15) appeared only in MGI and 14.3% (3) appeared only in UKB. The overlaps in phenome-wide significant hits across cohorts within weighting strategy are show as Venn diagrams in **Figure S10D-F**.

The unweighted meta-PheWAS identified 37 hits across 9 categories. The IP-weighted meta-PheWAS identified 22 hits across 5 categories. Of the 44 unique hits identified in both meta-PheWAS, 15.9% (7) appeared only in the IP-weighted meta-PheWAS. Notably, the IP-weighted meta-PheWAS identified a hit (NS_356.2: Aphasia and dysphasia) in a novel category (neurological). The overlaps in phenome-wide significant hits across weighting strategies are shown as Venn diagrams in **Figure S11**.

Of the 101 unique hits identified in any unweighted or PS-weighted PheWAS, 25.7% (n = 26) appeared only in PS-weighted PheWAS (**Figure S12**). PheWAS summary statistics are available in the **Supplementary Data File**.

### “Targeted” estimation of the sex-colorectal cancer log-odds ratio

The unweighted age-adjusted log-odds ratio for female sex and colorectal cancer were -0.098 (-0.164, -0.033), -0.164 (-0.247, -0.082), and -0.389 (-0.431, 0.348) for AOU, MGI, and UKB, respectively. Only the unweighted UKB estimate overlapped with the benchmark range of -0.414 to -0.271 based on 2018-2020 US SEER and UK^54^ estimates. The unweighted meta-analytic estimate was -0.284 (-0.316, -0.252). IP- and PS-weighting did not improve estimation in AOU, resulting in null estimates of -0.047 (-0.198, 0.104) and -0.084 (-0.191, 0.024), respectively. However, in MGI, weighting improved estimation with the IP-weighted confidence interval overlapping with (-0.217 (-0.419, -0.014)) and the PS-weighted point estimate falling within (-0.342 (-0.629, -0.056)) the benchmark range. IP-weighting did not change the UKB estimate (-0.398 (-0.461, -0.334)). The IP- and PS-weighted meta-analytic estimates (-0.335 (-0.392, -0.279) and -0.318 (-0.371, - 0.264), respectively) remained stable, driven by the UKB estimates. Along with unadjusted estimates, these results are shown in **Figure 5** and in **Table S3**.

**Figure 5.**
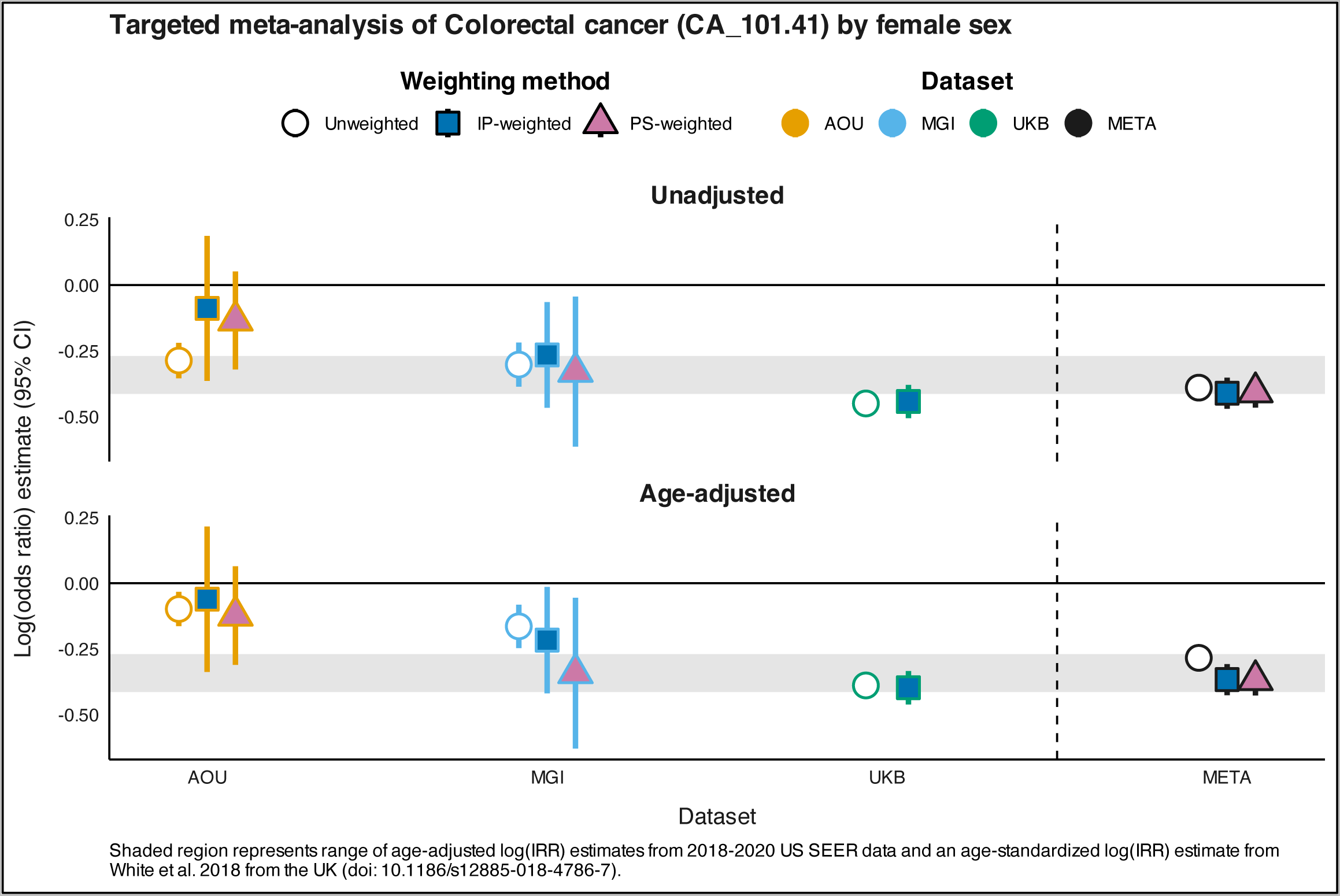
Within cohort and meta-analysis unadjusted and age-adjusted female log-odds ratio estimates (95% confidence interval) for colorectal cancer (phecode CA_101.41). Point estimate shapes and fill colors correspond to the weighting method (white circle, unweighted; dark blue square, inverse probability (IP)-weighted; pink triangle, poststratification (PS)-weighted). Line colors correspond to the cohort (orange, AOU; blue, MGI; green, UKB; black, meta-analysis). Shaded region represents range of age-adjusted log(incidence rate ratio [IRR]) estimates from 2018-2020 US SEER data and an age-standardized log(IRR) estimate from White et al. 2018 from the UK.^54^

## DISCUSSION

EHR-linked biobanks – such as AOU, MGI, and UKB analyzed here – are transforming the fields of epidemiology and health research. They offer valuable resources, comprising large longitudinal cohorts, with vast amounts of readily available structured and unstructured data and potential for data linkages at relatively low costs.^1,5–8,56^ However, the varying sampling mechanisms across these cohorts requires researchers to understand and address the impact of selection bias on various descriptive and inferential tasks (**Figure S1**).

We estimated IP- and PS-based selection weights for AOU and MGI and, along with previously described UKB IP-weights,^40^ evaluated their impact on common analyses currently undertaken in the field (impact on prediction is subject of forthcoming manuscript). Estimates of latent phenome dimensionality were marginally lower in cohorts with relatively higher phecode prevalences (e.g., AOU and MGI). The practical implication in terms of reduction in the denominator of a Bonferroni-corrected p-value from the number of total tests to the PCA-estimated number of independent tests would not have a meaningful impact.^57^ Further, p-value-identified results from untargeted hypothesis testing (as explored via a colorectal cancer PheWAS) for the strongest association signals remained largely unaltered following the introduction of selection weights. For example, the top 9 hits (and 12 total) from the unweighted meta-PheWAS were also identified in both weighted meta-PheWAS and the top 5 hits were the same in all meta-PheWAS (**Figure S11**). We also found that while weighting typically increases p-values, some p-values in MGI decreased, likely due to the presence of significant selection bias. Because of these results, we believe it is not crucial to use selection weights for exploring phenome structure and large-scale hypothesis testing tasks, particularly when such weights are not provided. If weights are readily available, the use of selection weights in this context is advisable. Significant hits from agnostic analyses should be followed by a targeted analysis where the importance of using weights is clearer.

For estimation tasks, like prevalence and effect size estimation, we recommend the use of selection weights to reduce potential selection bias. Regarding phecode prevalence estimation, we saw large changes in prevalence estimates after weighting (e.g., prevalence of MB_286.2: Major depressive disorder dropped 24 percentage points after IP-weighting in MGI) and these changes were phenome-wide (e.g., IP-weighted over unweighted MPR in AOU: 0.82). Regarding association estimation, we saw that the use of generic selection weights moved sex log-odds ratio estimates for colorectal to within the benchmark interval in MGI. However, in AOU estimates remained outside the benchmark interval even after weighting likely because of strong heterogeneity in the OR by race/ethnicity (**Figure S13**). When there is expected or known heterogeneity, stratified analyses are preferable, especially when the data are powered to do so (e.g., race/ethnicity-specific analyses in AOU). In such cases rational definition of the target population should be specific to race/ethnicity. In the case of targeted association estimation, we also recommend that weights be curated based on the outcome of interest, a conclusion supported by recent literature.^3,4,21^ Finally, *in all settings*, the use of selection weights is more important in samples that are less like the target population as in smaller and non-population-based cohorts like the MGI.

### Achievable goal is to reduce, not remove, bias

Weighted analyses are, historically, attempts to *remove* the impact of selection bias (e.g., on an association estimate) with respect to a defined target population.^25,41^ We developed selection weights based on explicit selection factors that were either publicly reported to have influenced recruitment strategies (as in AOU) or known to impact eligibility (as in MGI). However, these selection mechanisms are complex, and the true mechanisms are unknown. Thus, the use of selection weights is aimed at *reducing* bias rather than *removing* bias. This is particularly important in the case of Big Data where, while confidence intervals are narrow, effects of selection bias are not mitigated by increasingly large sample sizes.^58^ Additionally, some associations may be more or less prone to selection biases, but which associations are affected and how are not known. See **Section S2** for discussion on other weighting and EHR-related methodological concerns.

### Strengths and limitations

This study has multiple strengths. First, we utilized AOU and UKB data, which are large scale, public, and frequently used EHR-linked biobanks. Second, we utilized a range of methods to visualize and characterize EHR-linked biobanks. Third, we demonstrated estimation of IP- and PS-weights in AOU and provided code for recreating them. Fourth, the weights are based on NHIS data, a public resource with individual-level data representing a probabilistic sample of health conditions in the US population. Fifth, we used the new phecode X mapping table, which defines more phecodes compared to its predecessor (version 1.2), is built on ICD-10 data, and appears to have more accurate phecode definitions (an earlier version of this manuscript used phecode 1.2 mapping and found unexpected consequences of its phecode definitions; see **Section S3**).

However, our study also has several limitations. First, we cannot fully account for selection bias because the selection mechanisms are not fully known. Thus, our selection weights attempt to reduce – rather than remove – selection bias. Second, we do not know whether associations in weighted NHIS data can reliably replicate known associations in the US adult population. Third, while these cohorts are widely used, they vary noticeably in factors such as their geographical location, recruitment mechanisms, and access to EHR data (e.g., single medical system vs. primary care EHR). Future studies could examine more comparable cohorts to derive nuanced insights. Fourth, we conducted commonly performed meta-analyses, though the phenome has important sociobehavioral and environmental contributors that researchers need to consider when selecting cohorts for and interpreting meta-analyses. Finally, while our focus was on ways to lessen the impact of selection bias, there are multiple biases^59–61^ one needs to consider when conducting research and making causal inference. Future studies should investigate how these different biases affect data tasks and the role of method complexity in decision-making processes.

## CONCLUSION

We have introduced methods for assessing and comparing the effect of selection bias in EHR-linked biobanks and computed IP- and PS-weights for two US-based biobanks. These weights have the potential to reduce – not remove – selection bias as the selection mechanisms are not fully known. Our findings suggest that it is not crucial to use generic selection weights for exploring phenome structure (i.e., latent dimensionality, partial correlation across phecodes) and large-scale hypothesis testing. EHR-linked biobanks should provide detailed guidance on observation and recruitment processes and, where possible, make selection weights publicly available. Researchers should also clearly state their intended target population, the estimand of interest and describe recruitment and selection mechanisms from the source population. Systematic and rigorous exploration and comparisons of cohorts should be standard in analyses using multi-center EHR-linked biobank data.

## Data Availability

All of Us data are publicly available via researchallofus.org to those who fulfill their requirements.
Michigan Genomics Initiative data is available to researchers who receive access approvals from a University of Michigan Institutional Review Board.
UK Biobank data are available to researchers who receive approval (see ukbiobank.ac.uk).
Code used in these analyzes are publicly available at: https://github.com/maxsal/biobank_selection_weights.

## Acknowledgments

Michigan Genomics Initiative: The authors acknowledge the Michigan Genomics Initiative participants, Precision Health at the University of Michigan, the University of Michigan Medical School Central Biorepository, the University of Michigan Medical School Data Office for Clinical and Translational Research, and the University of Michigan Advanced Genomics Core for providing data and specimen storage, management, processing, and distribution services, and the Center for Statistical Genetics in the Department of Biostatistics at the School of Public Health for genotype data curation, imputation, and management in support of the research reported in this publication/grant application/presentation.

UK Biobank: This research has been conducted using the UK Biobank Resource under application number 88536.

All of Us: The All of Us Research Program is supported by the National Institutes of Health, Office of the Director: Regional Medical Centers: 1 OT2 OD026549; 1 OT2 OD026554; 1 OT2 OD026557; 1 OT2 OD026556; 1 OT2 OD026550; 1 OT2 OD 026552; 1 OT2 OD026553; 1 OT2 OD026548; 1 OT2 OD026551; 1 OT2 OD026555; IAA #: AOD 16037; Federally Qualified Health Centers: HHSN 263201600085U; Data and Research Center: 5 U2C OD023196; Biobank: 1 U24 OD023121; The Participant Center: U24 OD023176; Participant Technology Systems Center: 1 U24 OD023163; Communications and Engagement: 3 OT2 OD023205; 3 OT2 OD023206; and Community Partners: 1 OT2 OD025277; 3 OT2 OD025315; 1 OT2 OD025337; 1 OT2 OD025276. In addition, the All of Us Research Program would not be possible without the partnership of its participants.

