## Supplementary Materials for "To weight or not to weight? Studying the effect of selection bias in three large EHR-linked biobanks"

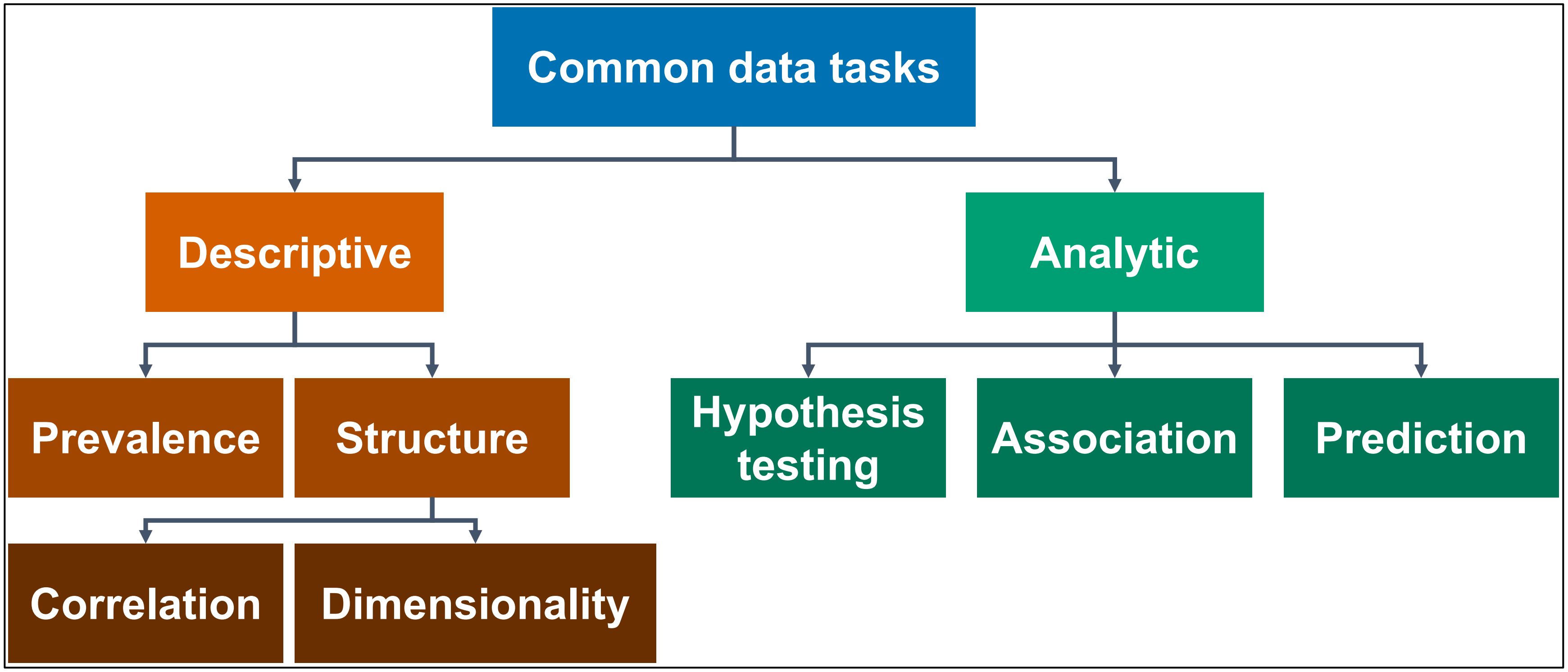


**Figure S1**. Flowchart depicting several common data tasks. This flowchart is subjective and not exhaustive.

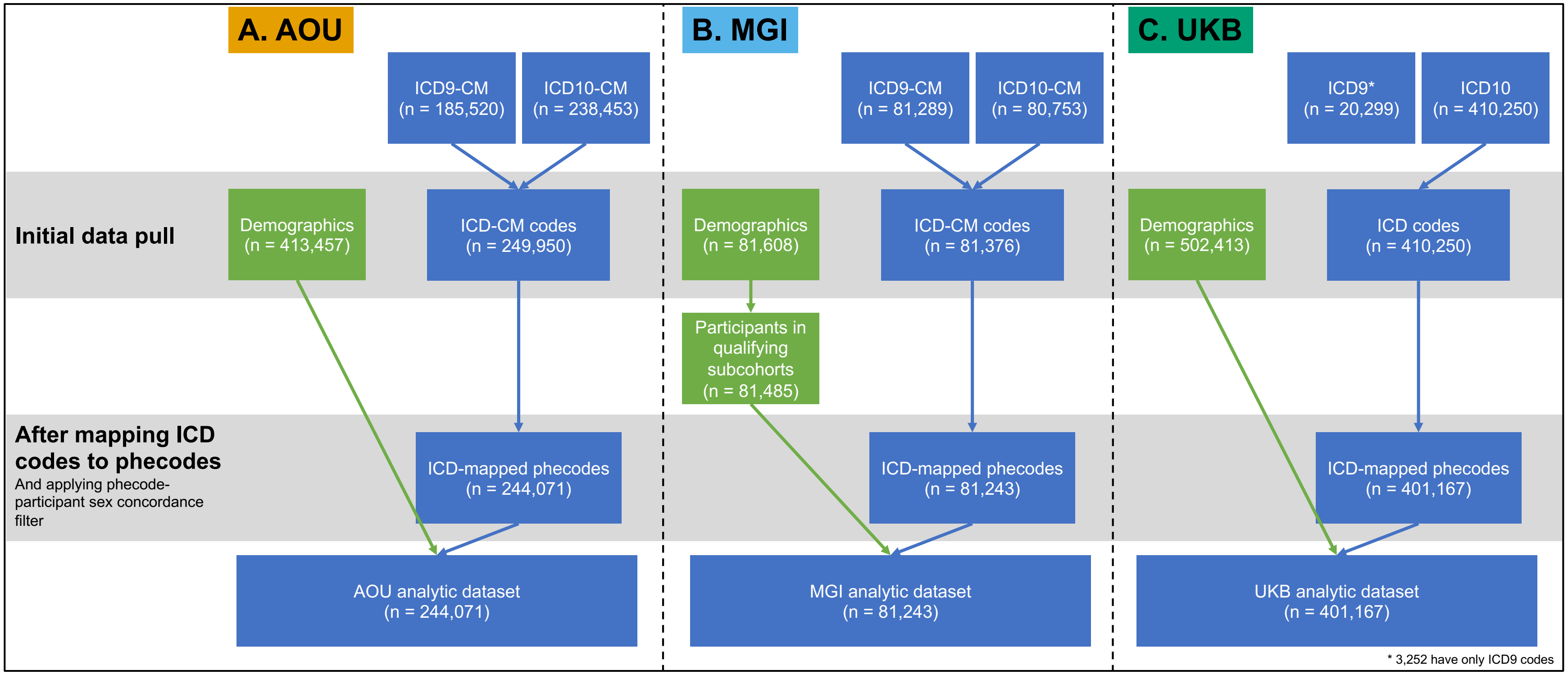


**Figure S2**. Flowcharts depicting samples sizes before and after filter and ICD-to-phecode mapping in AOU (panel A), MGI (panel B), and UKB (panel C).

**Table S1**. Phenotypes defined in paper and their qualifying phecode definitions

| Variable | Phecode | Description |
|---|---|---|
| Anxiety | MB_288 | Anxiety and anxiety disorders |
| Cancer | CA_100 | Malignant neoplasm of the head and neck |
| | CA_100.1 | Malignant neoplasm of the oral cavity |
| | CA_100.12 | Malignant neoplasm of the tongue |
| | CA_100.13 | Malignant neoplasm of the gums |
| | CA_100.14 | Malignant neoplasm of the floor of mouth |
| | CA_100.15 | Malignant neoplasm of the palate |
| | CA_100.2 | Malignant neoplasm of the oropharynx |
| | CA_100.3 | Malignant neoplasm of the nasopharynx |
| | CA_100.4 | Malignant neoplasm of the hypopharynx |
| | CA_100.5 | Malignant neoplasm of nasal cavities, middle ear, and accessory sinuses |
| | CA_100.6 | Malignant neoplasm of the larynx |
| | CA_100.7 | Malignant neoplasm of the pharynx |
| | CA_100.8 | Malignant neoplasm of the lip |
| | CA_100.9 | Malignant neoplasm of the salivary glands |
| | CA_101 | Malignant neoplasm of the digestive organs |
| | CA_101.1 | Malignant neoplasm of the esophagus |
| | CA_101.2 | Malignant neoplasm of stomach |
| | CA_101.21 | Malignant neoplasm of cardia |
| | CA_101.3 | Malignant neoplasm of the small intestine |
| | CA_101.4 | Malignant neoplasm of the lower GI tract |
| | CA_101.41 | Colorectal cancer |
| | CA_101.411 | Malignant neoplasm of colon |
| | CA_101.412 | Malignant neoplasm of appendix |
| | CA_101.42 | Malignant neoplasm of anus |
| | CA_101.6 | Malignant neoplasm of the liver and intrahepatic bile ducts |
| | CA_101.61 | Malignant neoplasm of the liver |
| | CA_101.62 | Malignant neoplasm of the intrahepatic bile ducts |
| | CA_101.7 | Malignant neoplasm of the gallbladder and extrahepatic bile ducts |
| | CA_101.71 | Malignant neoplasm of the gallbladder |
| | CA_101.8 | Malignant neoplasm of the pancreas |
| | CA_102 | Malignant neoplasm of the thoracic and respiratory organs |
| | CA_102.1 | Malignant neoplasm of the of bronchus and lung |
| | CA_102.3 | Malignant neoplasm of the trachea |
| | CA_102.5 | Malignant neoplasm of the heart, mediastinum, thymus, and pleura |
| | CA_102.51 | Malignant neoplasm of the heart |
| | CA_102.52 | Malignant neoplasm of the mediastinum |
| | CA_102.53 | Malignant neoplasm of the of pleura |
| | CA_102.54 | Malignant neoplasm of the thymus |
| | CA_103 | Malignant neoplasm of the skin |
| | CA_103.1 | Melanomas of skin |
| | CA_103.2 | Keratinocyte carcinoma |
| | CA_103.21 | Basal cell carcinoma |
| | CA_103.22 | Squamous cell carcinoma of the skin |
| | CA_103.3 | Carcinoma in situ of skin |
| | CA_104 | Malignant sarcoma-related cancers |
| | CA_104.1 | Malignant neoplam of the bone and/or cartilage |
| | CA_104.2 | Malignant neoplasm of retroperitoneum and peritoneum |
| | CA_104.3 | Malignant neoplasm of connective and soft tissue |
| | CA_104.4 | Malignant neoplasm of peripheral nerves* |
| | CA_104.5 | Gastrointestinal stromal tumor* |
| | CA_104.6 | Kaposi's sarcoma |

**Table S1**. Phenotypes defined in paper and their qualifying phecode definitions

| Variable | Phecode | Description |
|---|---|---|
| | CA_105 | Malignant neoplasm of the breast |
| | CA_105.1 | Malignant neoplasm of the breast, female |
| | CA_105.2 | Malignant neoplasm of the breast, male |
| | CA_106 | Gynecological malignant neoplasms |
| | CA_106.1 | Malignant neoplasm of external female genital organs and cervix |
| | CA_106.11 | Malignant neoplasm of the vulva |
| | CA_106.12 | Malignant neoplasm of the vagina |
| | CA_106.13 | Malignant neoplasm of the cervix |
| | CA_106.2 | Malignant neoplasm of the uterus |
| | CA_106.21 | Malignant neoplasm of endometrium |
| | CA_106.3 | Malignant neoplasm of the ovary |
| | CA_106.4 | Malignant neoplasm of the fallopian tube and uterine adnexa |
| | CA_106.6 | Malignant neoplasm of the placenta |
| | CA_107 | Malignant neoplasm of male genitalia |
| | CA_107.1 | Malignant neoplasm of the penis |
| | CA_107.2 | Malignant neoplasm of the prostate |
| | CA_107.3 | Malignant neoplasm of the testis |
| | CA_107.4 | Malignant neoplasm of epididymis |
| | CA_107.5 | Malignant neoplasm of spermatic cord |
| | CA_107.6 | Malignant neoplasm of the scrotum |
| | CA_108 | Malignant neoplasm of the urinary tract |
| | CA_108.4 | Malignant neoplasm of the kidney |
| | CA_108.41 | Malignant neoplasm of kidney, except pelvis |
| | CA_108.42 | Malignant neoplasm of renal pelvis |
| | CA_108.5 | Malignant neoplasm of the bladder |
| | CA_108.6 | Malignant neoplasm of urethra |
| | CA_108.7 | Malignant neoplasm of ureter |
| | CA_109 | Malignant neoplasm of the eye, brain and other parts of central nervous system |
| | CA_109.1 | Malignant neoplasm of eye |
| | CA_109.11 | Malignant neoplasm of orbit |
| | CA_109.12 | Malignant neoplasm of lacrimal gland and duct |
| | CA_109.13 | Malignant neoplasm of conjunctiva |
| | CA_109.14 | Malignant neoplasm of cornea |
| | CA_109.15 | Malignant neoplasm of retina |
| | CA_109.16 | Malignant neoplasm of choroid |
| | CA_109.2 | Malignant neoplasm of meninges |
| | CA_109.3 | Malignant neoplasm of brain |
| | CA_109.4 | Malignant neoplasm of spinal cord |
| | CA_109.5 | Malignant neoplasm of cranial nerve |
| | CA_110 | Malignant neoplasm of the endocrine glands |
| | CA_110.1 | Malignant neoplasm of the thyroid |
| | CA_110.3 | Malignant neoplasm of the parathyroid gland |
| | CA_110.4 | Malignant neoplasm of the pituitary gland and craniopharyngeal duct |
| | CA_110.5 | Malignant neoplasm of the pineal gland |
| | CA_112 | Malignant neoplasm of other and ill-defined sites |
| | CA_112.1 | Mesothelioma* |
| | CA_114 | Neuroendocrine tumors |
| | CA_114.1 | Malignant neuroendocrine tumors |
| | CA_114.11 | Exocrine pancreatic cancer |
| | CA_114.12 | Merkel cell carcinoma |
| | CA_114.2 | Benign neuroendocrine tumors |
| | CA_114.4 | Carcinoid tumors |
| | CA_114.41 | Intestinal carcinoid |

**Table S1**. Phenotypes defined in paper and their qualifying phecode definitions

| Variable | Phecode | Description |
|---|---|---|
| | CA_114.42 | Carcinoid tumor of the bronchus and lung |
| | CA_114.43 | Carcinoid tumor of the thymus |
| | CA_114.44 | Carcinoid tumor of the stomach |
| | CA_114.45 | Carcinoid tumor of the kidney |
| | CA_114.5 | Paraganglioma |
| | CA_114.6 | Pheochromocytoma |
| | CA_116 | Secondary malignant neoplasm |
| | CA_120 | Hemo onc - by cell of origin |
| | CA_120.1 | Myeloid |
| | CA_120.11 | Plasma cell |
| | CA_120.12 | Monocyte |
| | CA_120.13 | Erythroid |
| | CA_120.14 | Megakaryoblast |
| | CA_120.15 | Mast cell |
| | CA_120.2 | Lymphoid |
| | CA_120.21 | Mature B-cell |
| | CA_120.22 | Mature T-Cell |
| | CA_120.3 | Histocytoces |
| | CA_121 | Leukemia |
| | CA_121.1 | Acute leukemia |
| | CA_121.11 | Acute lymphoid leukemia |
| | CA_121.12 | Acute myeloid leukemia |
| | CA_121.2 | Chronic leukemia |
| | CA_121.21 | Chronic lymphoid leukemia |
| | CA_121.22 | Chronic myloid leukemia |
| | CA_121.23 | Chronic myelomonocytic (monocytic) leukemia |
| | CA_122 | Lymphoma |
| | CA_122.1 | Hodgkin lymphoma |
| | CA_122.11 | Nodular sclerosis Hodgkin lymphoma |
| | CA_122.2 | Non-Hodgkin lymphoma |
| | CA_122.21 | Follicular lymphoma |
| | CA_122.22 | Diffuse large B-cell lymphoma* |
| | CA_122.23 | Burkitt lymphoma |
| | CA_122.24 | T-cell lymphoma |
| | CA_122.25 | Anaplastic large cell lymphoma |
| | CA_122.26 | Extranodal NK/T-cell lymphoma, nasal type* |
| | CA_123 | Multiple myeloma and malignant plasma cell neoplasms |
| | CA_123.1 | Multiple myeloma |
| | CA_124 | Myeloproliferative disorder |
| | CA_124.3 | Polycythemia vera |
| | CA_124.5 | Essential thrombocythemia |
| | CA_124.6 | Myelodysplastic syndrome |
| | CA_124.7 | Chronic myeloproliferative disease* |
| | CA_124.8 | Myelofibrosis |
| | CA_125 | Other malignant neoplasms of lymphoid, hematopoietic and related tissue |
| | CA_128 | Estrogen receptor status |
| | CA_128.1 | Estrogen receptor postitive status [ER+] |
| | CA_128.2 | Estrogen receptor negative status [ER-] |
| | CA_130 | Cancer (solid tumor, excluding BCC) |
| | CA_132 | Sequelae of cancer |
| Coronary artery disease | CV_404.2 | Coronary atherosclerosis [Atherosclerotic heart disease] |
| Depression | MB_286.2 | Major depressive disorder |

**Table S1**. Phenotypes defined in paper and their qualifying phecode definitions

| Variable | Phecode | Description |
| --- | --- | --- |
| Diabetes | EM_202 | Diabetes mellitus |

Visit https://phewascatalog.org (phecodeX) and https://github.com/PheWAS/PhecodeX

**Table S2**. Definition of variables by cohort used throughout paper

| | AOU | MGI | UKB | NHIS (2019)* |
|---|---|---|---|---|
| Age | Age at last diagnosis | Age at last diagnosis | Age at consent: date of consent (field ID 200) minus date of birth (field IDs 34, 52) | Age at screening (AGEP_A) |
| Sex | Self-reported sex at birth (field name: sex_at_birth_concept_id) | Self-report EHR | Acquired by central registry at recruitment, may be updated by individual (field ID 31) | SEX_A |
| Race/ethnicity | Self-reported race ethnicity (field names: race_source_concept_id, ethnicity_source_concept_id) | Self-report EHR | Self-report survey (field ID 21000) | HISPALLP_A |
| BMI | Median of EHR values | Median of EHR values | Median of assessed values (field ID 21001) | BMICAT_A (HEIGHTTC_A, WEIGHTLBTC_A) |
| Smoking status | Self-report (concept IDs: 1585857, 1585860) | Self-report EHR | Survey (field ID 20116) | SMKCIGST_A |
| Anxiety | Phecode MB_288: Anxiety and anxiety disorders | | | GADCAT_A |
| Cancer | See Table S1 | | | CANEV_A |
| Coronary artery disease | Phecode CV_404.2: Coronary atherosclerosis [Atherosclerotic heart disease] | | | CHDEV_A |
| Depression | Phecode MB_286.2: Major depressive disorder | | | PHQCAT_A |
| Diabetes | Phecode EM_202: Diabetes mellitus | | | DIBEV_A |

* visit https://www.cdc.gov/nchs/nhis/2019nhis.htm for more information

### Supplementary Methods

*Inverse probability weighting*

In MGI, we estimated the first term, $P(S_{external} = 1|\boldsymbol{X})$, by fitting a simplex regression model for the known design probabilities using NHIS data. We estimated the numerator of the second term, $P(S = 1|\boldsymbol{X}, S_{all} = 1)$, using a logistic regression model. We considered the set of selection factors, $\boldsymbol{X}$: age (≥ 50 indicator), female sex, BMI (categorical), non-Hispanic White race/ethnicity, and EHR-derived binary indicators for anxiety, depression, diabetes, cancer, and hypertension (variable definitions in **Table S2**). Cancer was not included directly in the estimation procedure above because the small prevalence of cancer in NHIS led to unstable model fitting.[1] Instead, a cancer factor, $\gamma_{cancer}$, defined as $\frac{P(\text{Cancer}|\boldsymbol{X},S=1)}{P(\text{Cancer}|\boldsymbol{X})}$, was estimated by fitting logistic regression models with the same $\boldsymbol{X}$. The probabilities, $\psi$, were multiplied by this factor (i.e., $\psi\gamma_{cancer}$).

In AOU, we flexibly selected $\boldsymbol{X}$ by splitting the data in half and fitting a lasso-penalized logistic regression model on $\boldsymbol{X}$ and all possible pairwise interactions using the glmnet R package (version 4.1-8). We considered a set of selection factors, $\boldsymbol{X}$: age ((≥ 50 indicator), female sex, non-Hispanic White race/ethnicity, non-heterosexual orientation (yes/no), health insurance coverage status (yes/no), annual household/family income (≥ $75,000), educational attainment (at least high school graduate or equivalent), and region of residence (indicators for West, South, and Northeast) (variable definitions in **Table S2**). Using 10-fold cross-validation, we selected the largest $\lambda$ such that the error is within 1 standard error of the minimum to result in a parsimonious model. Of the 55 possible main effect and interaction terms, 39 were selected by this model (**Table S3**) and (along with the main effect for West region) were then used as the final set of $\boldsymbol{X}$ to estimate IP weights in the other half of the data as described for MGI above. The indicator variables for income, health insurance status, and non-Hispanic White race/ethnicity were the three most important variables (**Figure S2**). In both cohorts, the resulting probabilities were winsorized at the 2.5$^{th}$ and 97.5$^{th}$ percentiles.

We note that augmented inverse probability weighting (AIPW) is a doubly robust weighting method that may be of interest to the reader; see [2–4].

*Poststratification weighting*

In AOU, we considered the set of $\boldsymbol{X}$: age (≥ 50 indicator), female sex, non-Hispanic White race/ethnicity, sexual orientation (non-heterosexual indicator), health insurance coverage status (yes/no), annual household/family income (≥ $75,000 indicator), and region of residence (categorical). In MGI we considered the set of selection factors, $\boldsymbol{X}$: age (≥ 50 indicator), female sex, non-Hispanic White race/ethnicity, BMI (categorical), smoking status (ever/never), and EHR-derived history of anxiety, cancer, depression, diabetes, and hypertension.

We note that other there are other weighting methods relying only on summary statistics like calibration, raking, and pseudo likelihood that may be of interest to the reader; see [5–7].

*Correlations*
We also explored the correlation structure of unweighted and weighted phenomes through partial correlations. Unweighted partial correlations were calculated between pairs of traits, $X$ and $Y$, adjusted for age and sex, using the ppcor R package (version 1.1). [8] Weighted partial correlations were approximated as the coefficient $\beta_X$ from the weighted multiple linear regression model $Y = \beta_0 + \beta_X X + \boldsymbol{\beta_Z Z}$, where $X$, $Y$, and $\boldsymbol{Z}$ were mean standardized and $\boldsymbol{Z}$ were age and female sex. For $X, Y$ pairs where one trait was sex-specific, the other trait was limited to individuals of that sex, and sex was not included as a covariate. Network graphs of correlations with absolute values greater than 0.3 were constructed to visually inspect the structure. All traits were treated as binary based on the presence of a single phecode in the EHR. (See **Section S1** for results).

*PheWAS*
The data were prepared as described in Salvatore and colleagues [9] at the one-year prior to colorectal cancer diagnosis threshold. For sex-specific phecodes, those with discordant sex were treated as missing. (Of note, some ICD codes do not map to phecodes). Logistic regression models were fit as follows:

$$logit\left(P(\mathrm{CA}_{101.41} = 1 | k, \boldsymbol{covariates})\right) = \beta_0 + \beta_k k + \boldsymbol{\beta}_{\mathbf{covariates}}\mathbf{covariates} \qquad \textit{Eq. (S1)}$$

where CA_101.41 (the phecode for colorectal cancer) is an indicator for the outcome, $k$ represents the exposure phecode $k$ (indicator), and covariates are age at one-year prior to colorectal cancer diagnosis (continuous), female sex (indicator), and length of EHR follow-up (continuous).

Phenomewide significant hits were identified using a conservative multiple testing corrected threshold of 0.05 divided by the number of *total* tests. Weighted logistic regression models were fit using svyglm from the survey R package.[10] In cases where a given exposure phecode did not have both (1) at least 20 occurrences and (2) at least 10 individuals with the exposure and colorectal cancer, weighted Firth bias-corrected logistic regression (logistf R package version 1.26.0) was used to address concerns about separation.

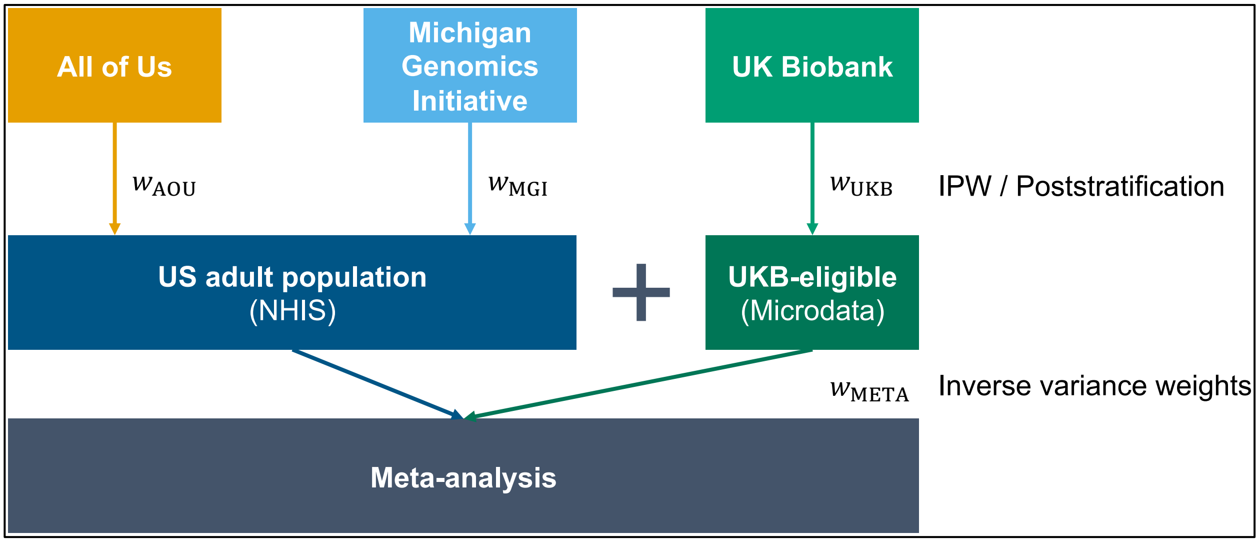


**Figure S3**. A schematic representation of the targeted and untargeted association analyses pipelines carried out in the manuscript.

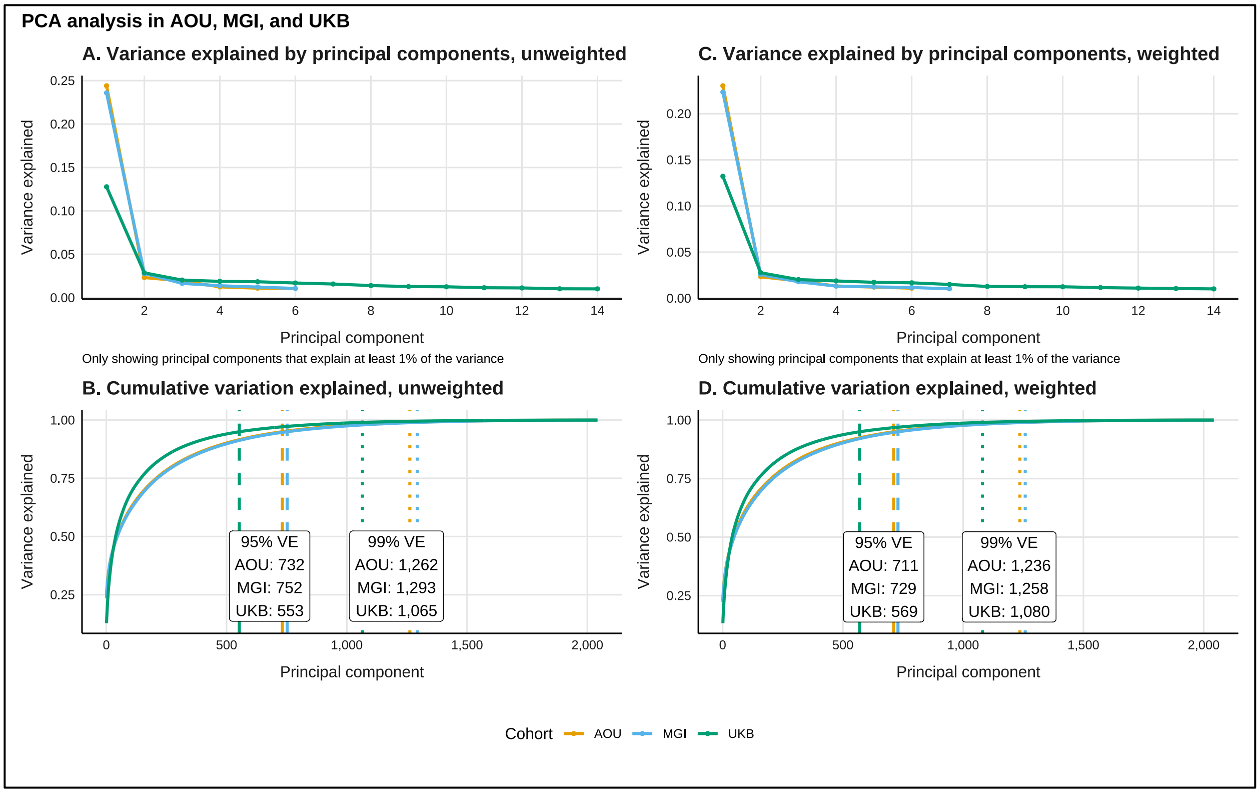


**Figure S4**. Principal components (PC) analysis in All of Us (AOU), the Michigan Genomics Initiative (MGI), and the UK Biobank (UKB). Panel A shows all principal components explain at least 1% of variation. Panel B shows the cumulative proportion of variance explained (VE) and reports variance explanation thresholds. The vertical dashed lines represent the number of PCs that explain at least 95% of total variance. The vertical dotted lines represent the number of PCs that explain at least 99% of the total variance.

**Section S1. Unweighted and weighted partial correlations**

Network diagrams depicting unweighted and weighted partial correlation coefficients with absolute values greater than 0.3 (an arbitrary threshold) in AOU is shown in **Figure S5** (MGI and UKB shown in **Figures S6** and **S7**). We can see clusters of correlated traits within endocrine/metabolic and musculoskeletal categories, as well as a cluster including both digestive and neurological traits. A small reduction in correlations with absolute values greater than 0.3 were observed after weighting (2,533 vs. 2,474). Interestingly, we see strong correlations with neoplasm traits in MGI (**Figure S6**), which largely disappear after weighting. There are distinct clusters within musculoskeletal traits and across circulatory system and endocrine/metabolic traits in UKB, which remain after weighting. The number of strong (absolute value > 0.3) correlations in UKB slightly increases after weighting (1,674 vs 1,757). **Figures S8** and **S9** depict the distribution of the unweighted and weighted partial correlation coefficients in each cohort, respectively. Generally, correlations tend to be highest in MGI followed by AOU and then UKB. Comparing the two US-based cohorts, AOU (**Figure S5**) and MGI (**Figure S6**), we see that, while the prevalences of traits involved in these networks are comparable, the network in MGI is denser compared to AOU.

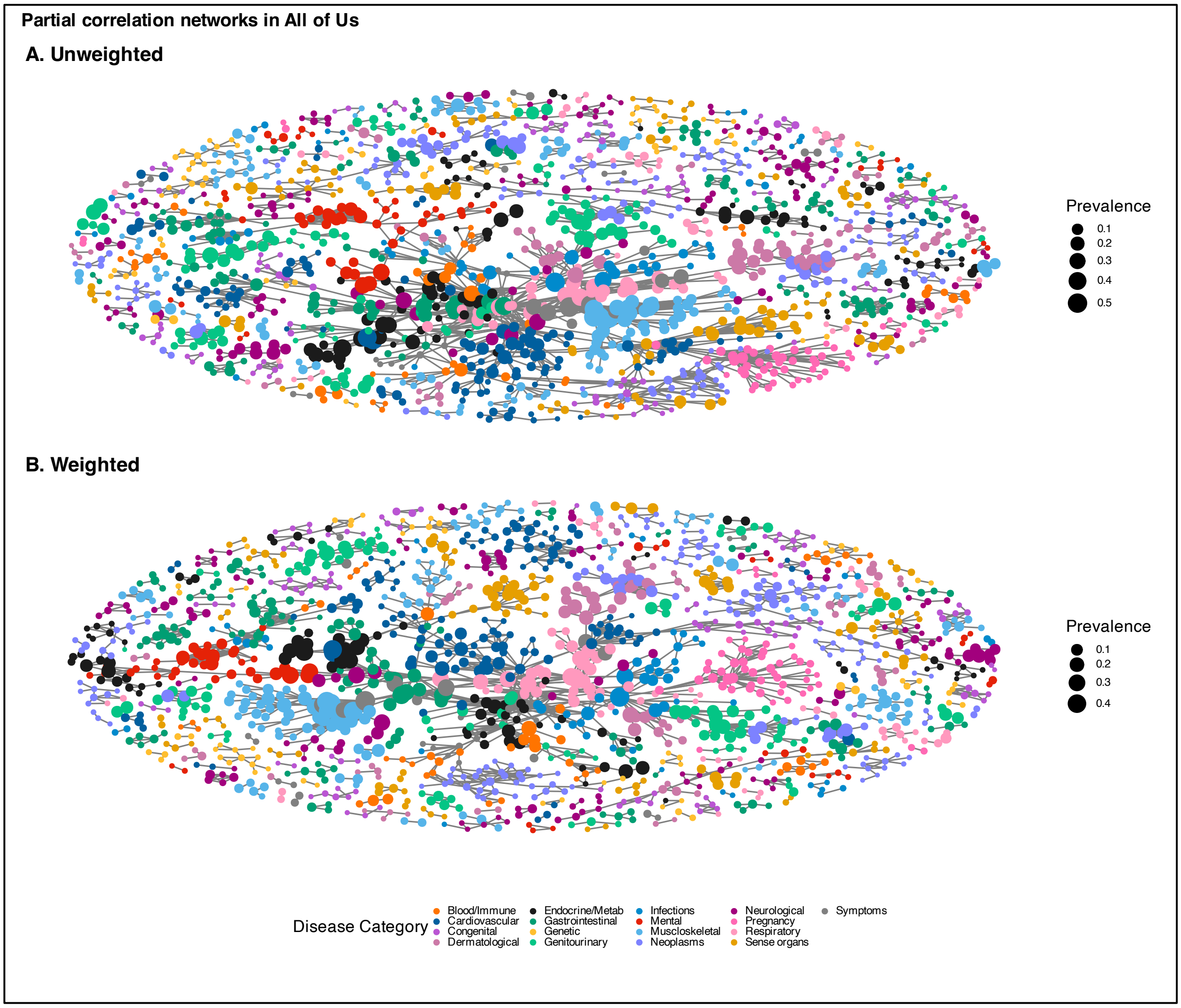


**Figure S5**. Unweighted (panel A) and inverse probability-weighted (panel B) network plots of the partial correlation structure of medical phenomes in All of Us. Correlation coefficients are adjusted for age and sex. Only correlations with an absolute value greater than or equal to 0.3 are shown. The size of the nodes corresponds to the prevalence of the trait in its cohort and the color corresponds to the phecode category. Corresponding figures for MGI and UKB are in **Figures S6** and **S7**, respectively.

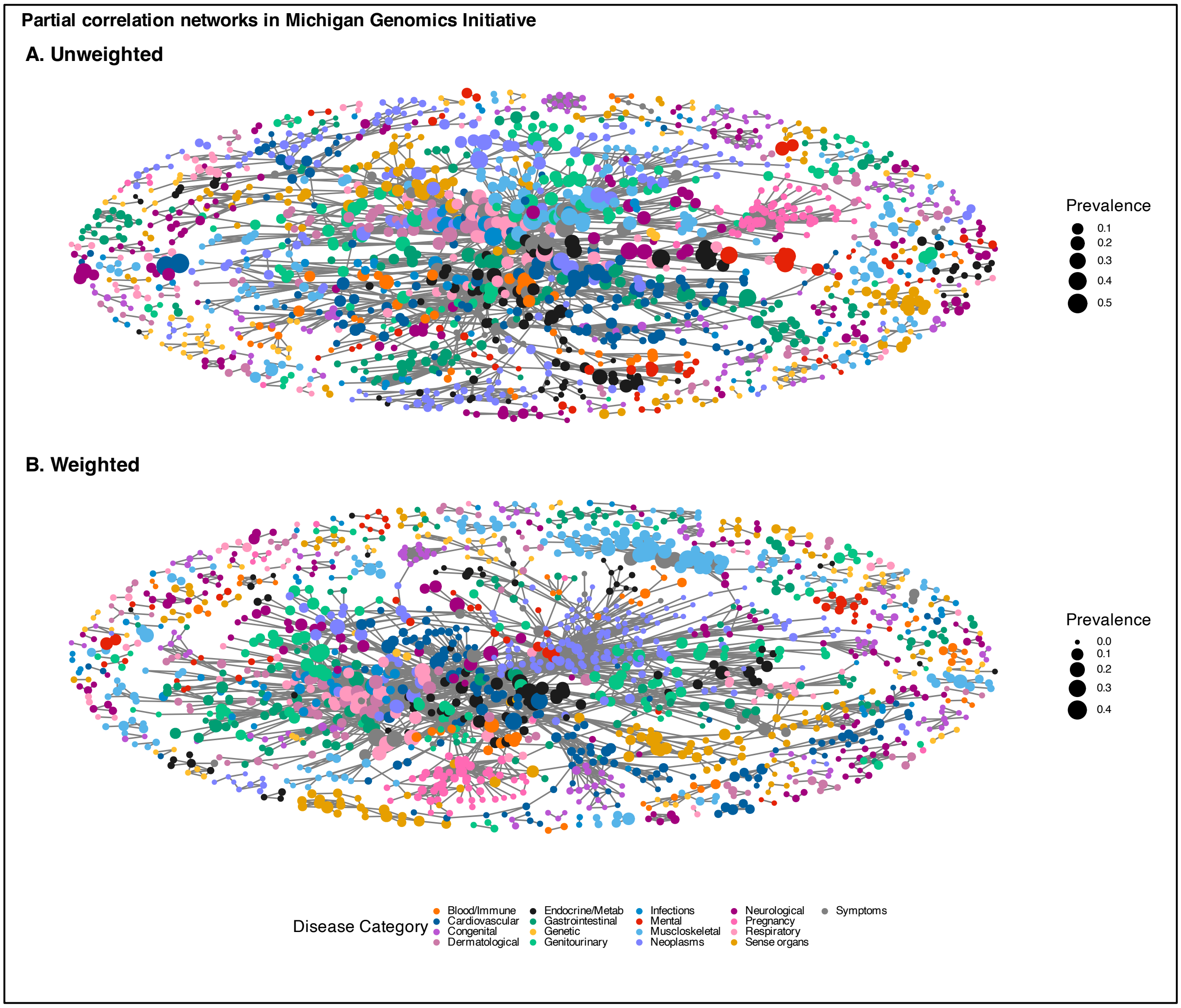


**Figure S6**. Unweighted (panel A) and inverse probability-weighted (panel B) network plots of the partial correlation structure of medical phenomes in MGI. Correlation coefficients are adjusted for age and sex. Only correlations with an absolute value greater than or equal to 0.3 are shown. The size of the nodes corresponds to the prevalence of the trait in its cohort and the color corresponds to the phecode category. Corresponding figures for AOU and UKB are in **Figures S5** and **S7**, respectively.

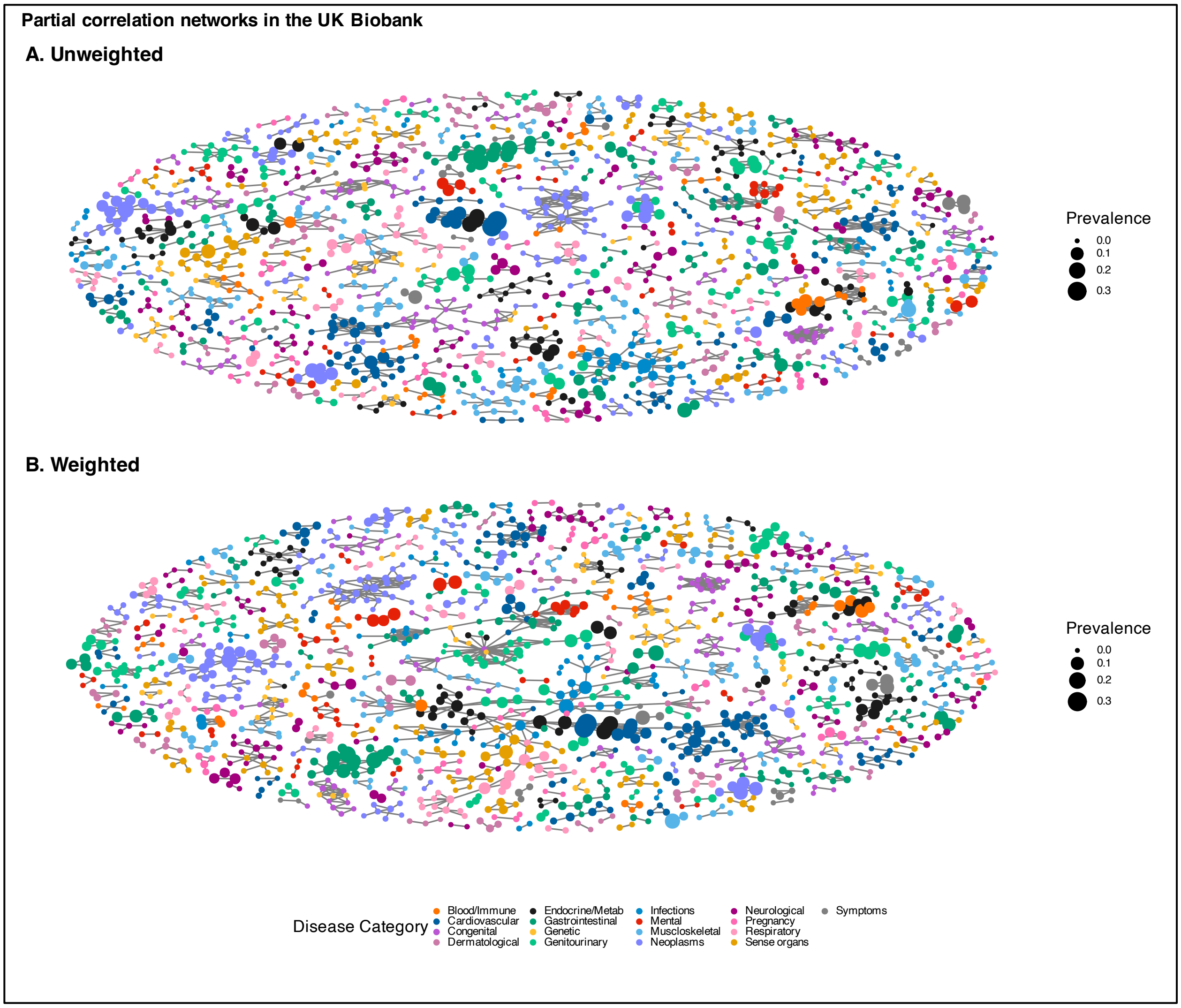


**Figure S7**. Unweighted (panel A) and inverse probability-weighted (panel B) network plots of the partial correlation structure of medical phenomes in UKB. Correlation coefficients are adjusted for age and sex. Only correlations with an absolute value greater than or equal to 0.3 are shown. The size of the nodes corresponds to the prevalence of the trait in its cohort and the color corresponds to the phecode category. Corresponding figures for AOU and MGI are in **Figures S5** and **S6**, respectively.

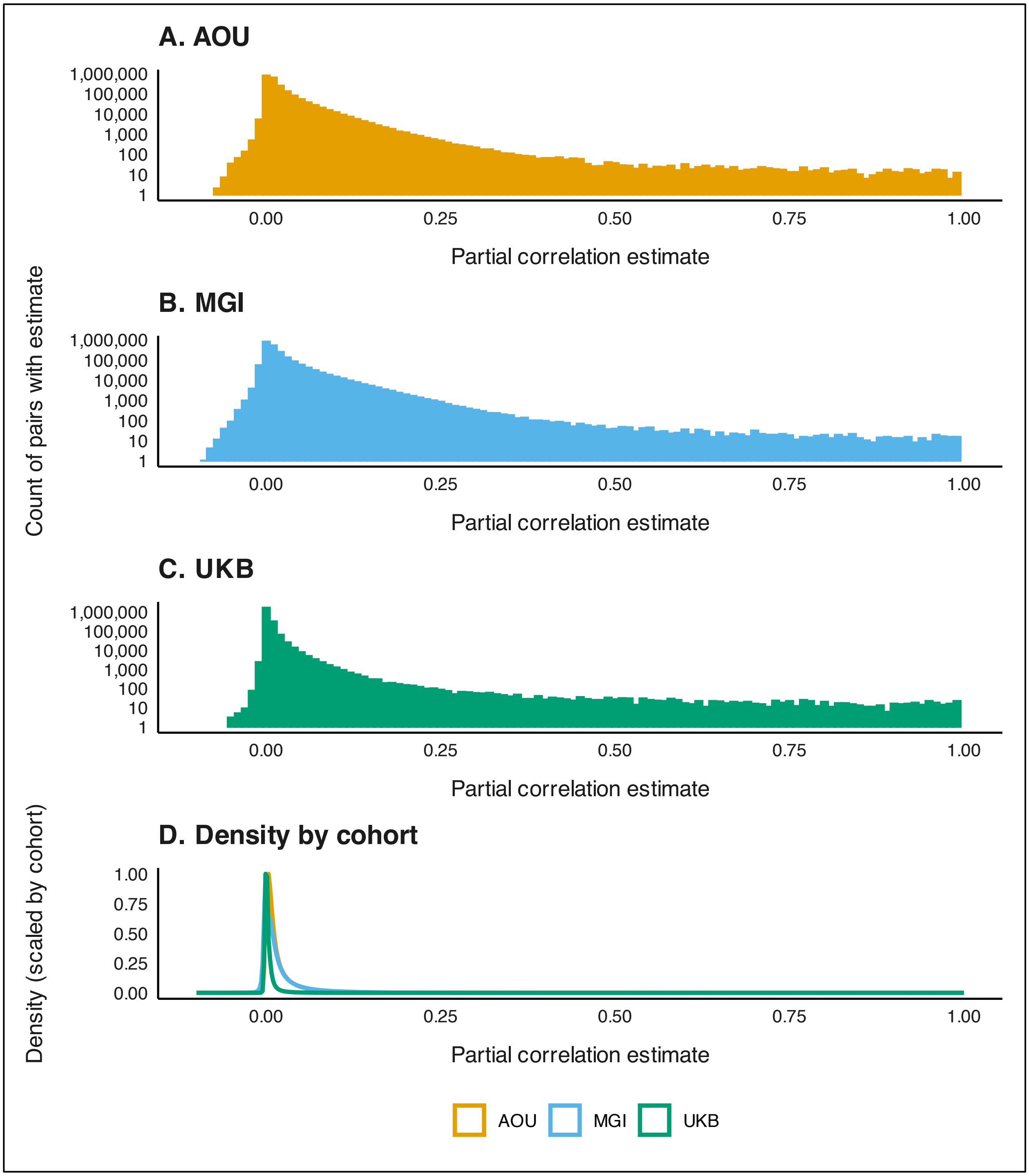


**Figure S8**. Distribution of unweighted partial correlations across medical phenomes. Partial correlations were adjusted for age and, if both codes in the pair applied to both sexes, sex.

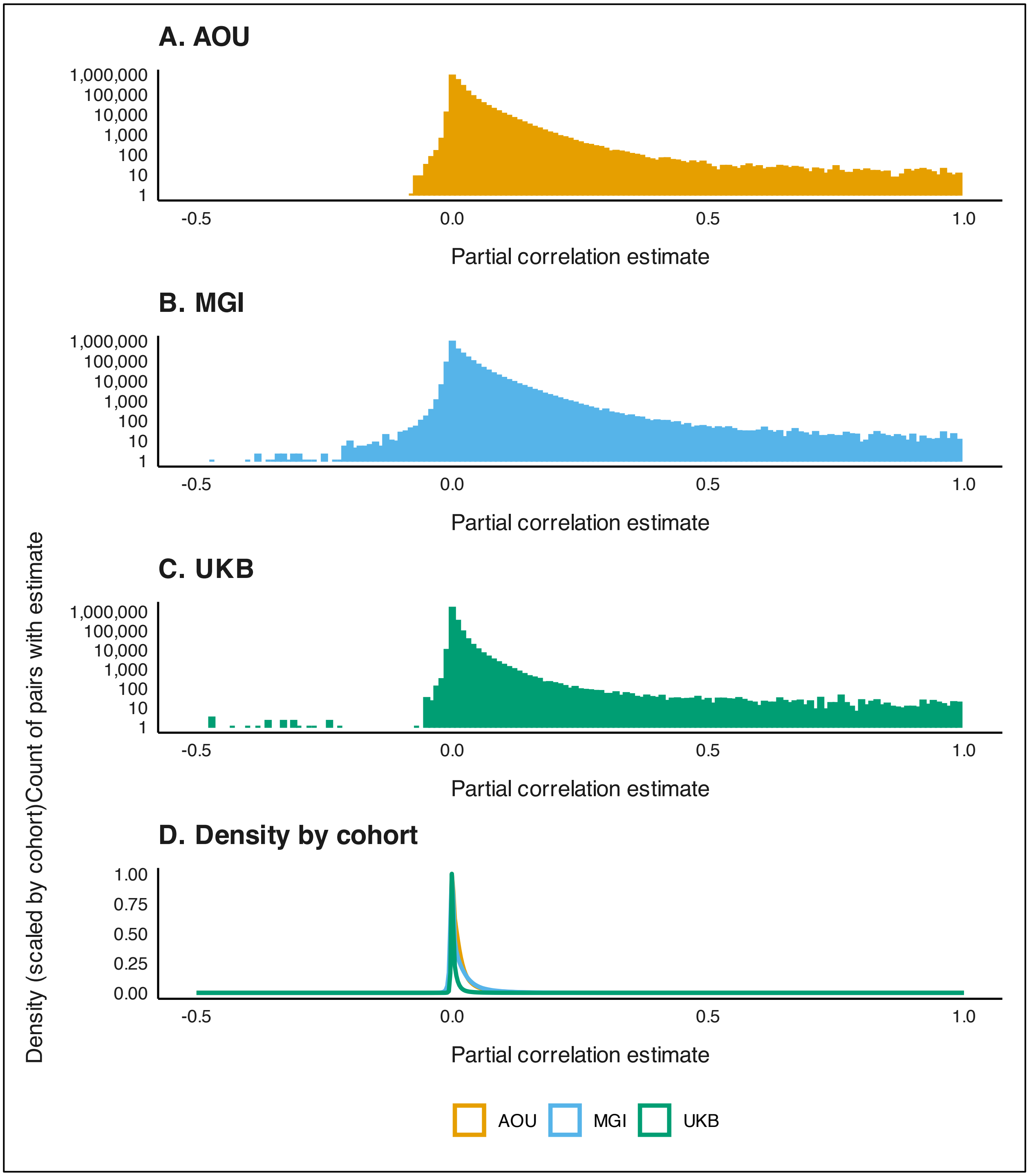


**Figure S9**. Distribution of weighted partial correlations across medical phenomes. Partial correlations were adjusted for age and, if both codes in the pair applied to both sexes, sex. IP-based weights were used for AOU and MGI and IP-based weighted developed by van Alten and colleagues[11] were used for UKB.

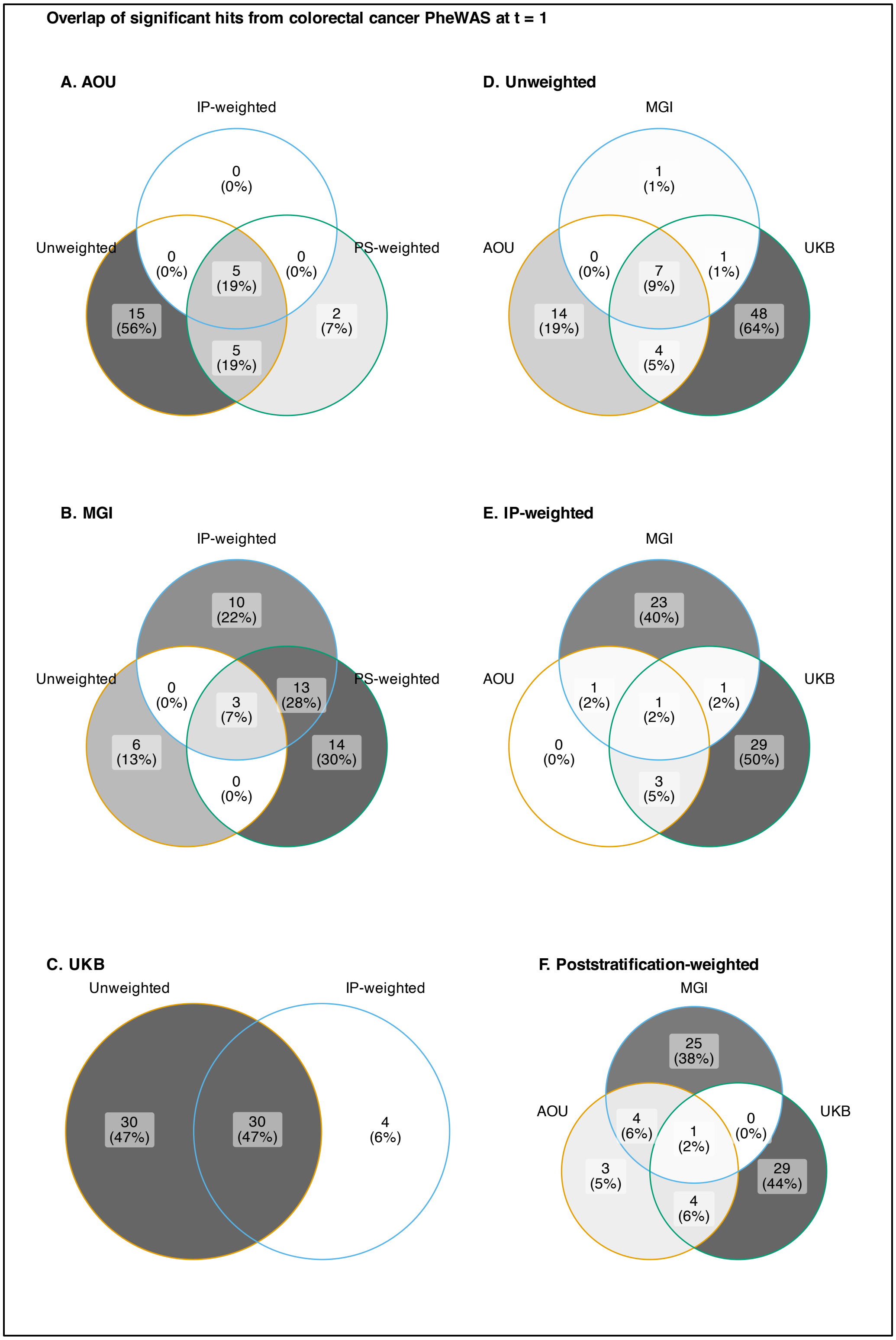


**Figure S10**. Venn diagrams comparing the overlap in phenome-wide significant hits from unweighted and weighted colorectal cancer PheWAS in AOU, MGI, and UKB.

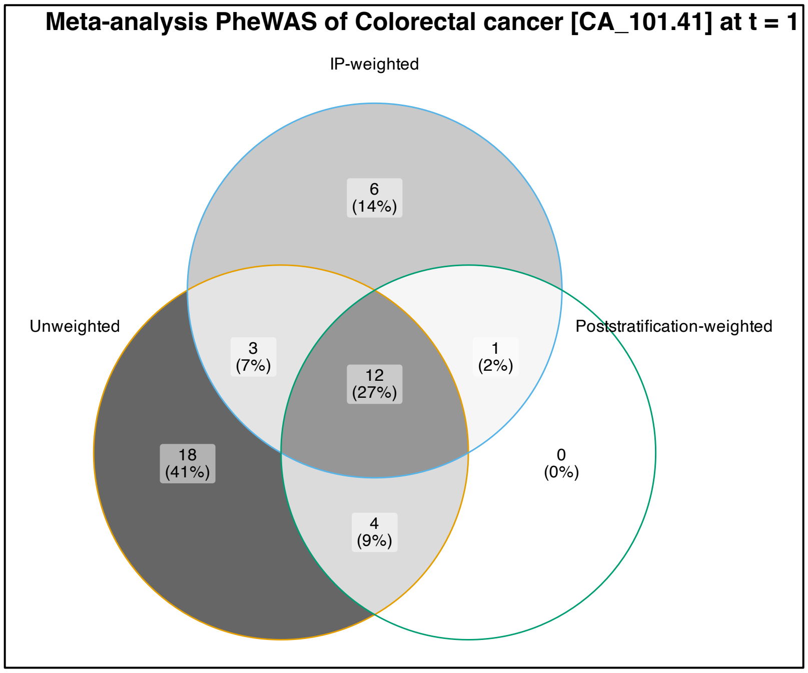


**Figure S11**. Venn diagrams comparing the overlap in phenome-wide significant hits from meta-analysis PheWAS.

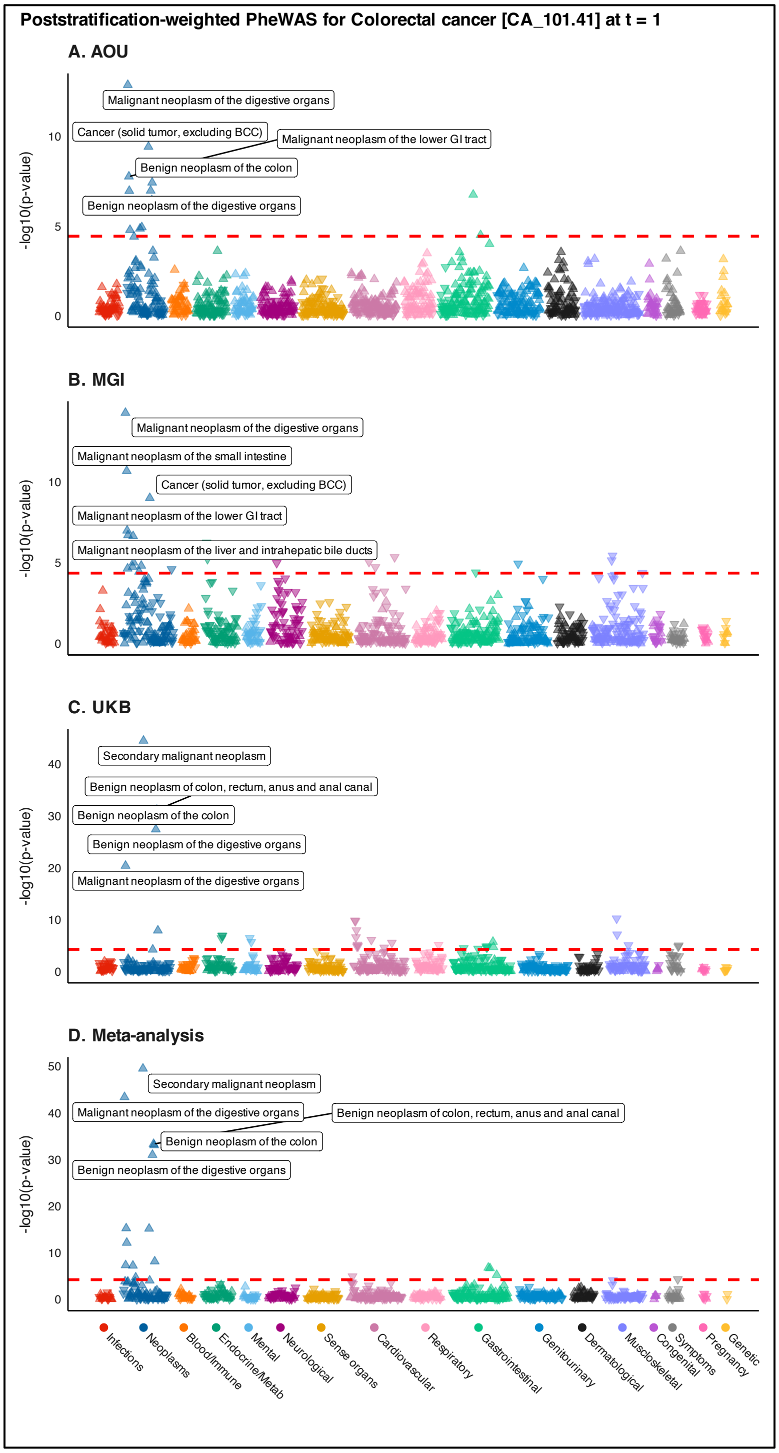


**Figure S12**. Manhattan plots summarizing poststratification-weight (panels A-D) phenomewide association studies for colorectal cancer in All of Us and the Michigan Genomics Initiative and the inverse probability weighted UK Biobank using 1:2 case:non-case matched data restricted to one year prior to initial diagnosis along with the corresponding meta-analsysis. The dashed red line represents the Bonferroni-corrected p-value threshold (-log10(0.05/number of traits)). The five traits with the smallest p-values are labeled. The upward (downward) orientation of the triangle indicates a positive (negative) association. Plots corresponding to unweighted and IP-weighted PheWAS are presented in **Figure 4**.

**Table S3**. Female log odds ratio estimate (95% confidence interval) for colorectal cancer (phecode CA_101.41).

| Weighting | Covariates | AOU | MGI | UKB | META |
|---|---|---|---|---|---|
| Unweighted | None | **-0.287** (-0.354, -0.220) | **-0.303** (-0.387, -0.219) | **-0.450** (-0.492, -0.409) | **-0.390** (-0.423, -0.358) |
| | Age | **-0.098** (-0.164, -0.033) | **-0.164** (-0.247, -0.082) | **-0.389** (-0.431, -0.348) | **-0.284** (-0.316, -0.252) |
| IP-weighted | None | -0.037 (-0.188, 0.113) | **-0.266** (-0.467, -0.065) | **-0.443** (-0.506, -0.380) | **-0.373** (-0.429, -0.317) |
| | Age | -0.047 (-0.198, 0.104) | **-0.217** (-0.419, -0.014) | **-0.398** (-0.461, -0.334) | **-0.335** (-0.392, -0.279) |
| PS-weighted | None | -0.135 (-0.321, 0.052) | **-0.329** (-0.615, -0.044) | **-0.443** (-0.506, -0.380) | **-0.408** (-0.466, -0.349) |
| | Age | -0.123 (-0.311, 0.064) | **-0.342** (-0.629, -0.056) | **-0.398** (-0.461, -0.334) | **-0.368** (-0.427, -0.310) |

* Meta-analysis results include IP-weighted estimate from UKB

Abbrevs: AOU, All of Us; IP, inverse probability; META, meta-analysis; MGI, Michigan Genomics Initiative; PS, poststratification; UKB, UK Biobank

Bolded point estimates are statistically significant at the 95% confidence level

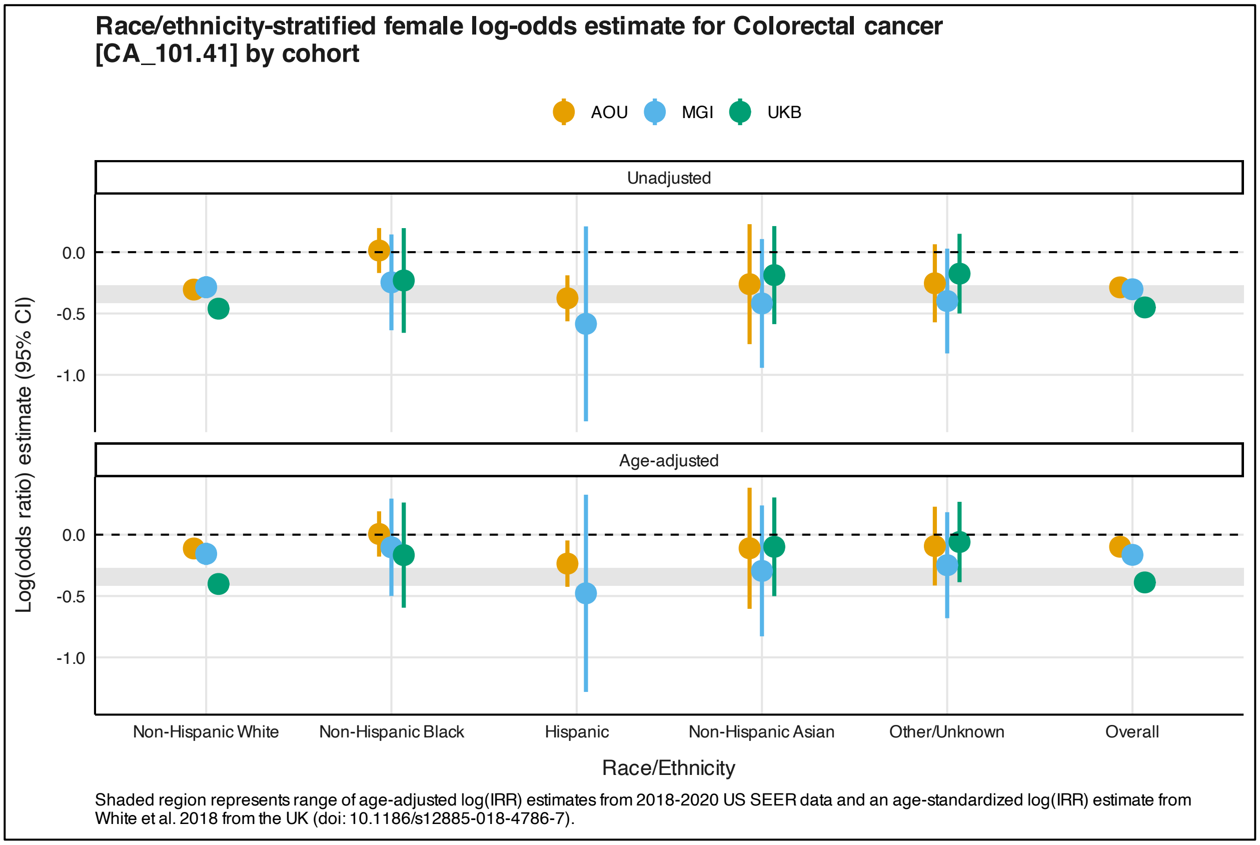


**Figure S13**. Unadjusted and age-adjusted female log-odds ratio estimate (95% confidence interval) for colorectal cancer (phecode CA_101.41) by race/ethnicity and cohort. Point estimate shapes correspond to the weighting method (circle, unweighted; square, IP-weighted; triangle, PS-weighted). Line colors correspond to the cohort (orange, AOU; blue, MGI; green, UKB). Shaded region represents range of age-adjusted log(incidence rate ratio [IRR]) estimates from 2018-2020 US SEER data and an age-standardized log(IRR) estimate from White et al. 2018 from the UK.[12]

### Section S2. Comments on methodological considerations in EHR-based data analysis

Weighting-based analytic approaches present a relatively simple way for researchers to improve the generalizability of their results and help *reduce* (not *remove*) selection bias. IP weights are preferred to PS weights though they rely on the assumption that the weighting model is correctly specified. Regression-based weights can be made more flexible through the use of indicator variables (as in our AOU IP-weights and in van Alten and colleagues[13]), though non-parametric methods like random forest can be used. When individual-level data from the target population is not available, PS weights can be estimated using summary-level strata probabilities (provided these probabilities are conditionally independent). When selection weights are unavailable, methods like covariate or propensity score adjustment, which are simple to implement, can be considered to address in some situations where selection bias is a concern.

Beyond introductory papers,[14–19] substantial work has focused specifically on traditional methodological concerns including confounding,[20,21] misclassification,[7,22,23] missing data,[24–30] and selection bias and cohort representativeness[1,6,7,31–34] related to EHR-based cohorts. For example, traits defined using the phecode framework have demonstrated reduced misclassification compared to ICD codes.[35] One method to further reduce the impact of misclassification, described by Hubbard and colleagues, relies on EHR-derived probabilistic phenotyping.[22] Others have described methods using manual chart review on a subset of data to improve EHR-derived phenotypes.[23,36,37] Beesley and Mukherjee developed three novel likelihood-based bias correction strategies to address outcome misclassification of EHR-derived disease status.[7] Teixeira and colleagues explored incorporation of unstructured data like doctors notes, which improved the identification of hypertensive individuals compared to using ICD codes and blood pressure reading cutoffs alone.[38] Missing data is another issue that has received attention to avoid loss of power and inducing selection bias (via complete case analyses) and aid in meeting assumptions necessary for multiple imputation.[25] One avenue is using non-missing genotype data available in EHR-linked biobanks to inform imputation, which demonstrated improvements in imputation of cardiovascular related measurements.[39] This idea could be extended using exposure polygenic risk scores[40] to inform imputation of missing exposure data.

One consideration broadly applicable in health research but is particularly acute in EHR-based analyses is *target validity*. Westreich and colleagues have defined this as a joint measure of internal and external validity of an effect estimate with respect to a specific target population.[41] Historically, internal validity, the notion that an estimate reflects the true underlying parameter in the study population, has taken precedence over external validity, that the parameter in the study population is representative of the true parameter in the target population. However, because of observation mechanisms and recruitment strategies into EHR-linked biobanks, the target population is almost certainly never (1) exactly the study sample or (2) the population of which the study sample is a simple random sample.[41] EHR researchers should think critically regarding who the results are intended for or representative of before beginning an analysis and make their target populations explicit in their work. We believe it is critical for researchers to consider weighted approaches that account for both the observation and recruitment mechanisms in each cohort (including potential subcohorts) and differences in the distribution of key characteristics between the analytic cohort and the target population.

We want to highlight some considerations that are hallmarks of EHR analysis. One such consideration is *informed presence*, defined by Goldstein and colleagues as "the notion that inclusion in an EHR is not random but rather indicates that the subject is ill, making people in EHRs systematically different from those not in EHRs."[42] This resulting discrepancy harms generalizability to general populations who tend to be healthier than those in the EHR data sample and results in bias. This concept extends to individuals within the EHR – those that are sicker tend to have more encounters and records than those who are healthier – and, in some cases, to records in the EHR (e.g., lab results). This phenomenon is illustrated by Agniel and colleagues, which shows that the presence and timing of laboratory results was more informative than the value of the laboratory results themselves. [43] Interested readers can learn more about informed presence elsewhere. [14,42,44–46] Including EHR metadata, like length of follow-up, number of encounters, density of laboratory measurements, and visit type (e.g., outpatient vs inpatient vs emergency), and careful selection or matching of controls in analyses are recommended to improve exchangeability and attempt to make EHR observation mechanisms comparable.

### Section S3. Investigation into infectious diseases peak in AOU PheWAS using phecode 1.2 mapping tables

An earlier version of the manuscript was performed using the phecode 1.2 mapping tables instead of phecode X. The Manhattan plot representing the colorectal cancer PheWAS in AOU in **Figure S14A** shosw a peak in the infectious disease category. The top hit is Human immunodeficiency virus [HIV] disease, or phecode 071 in the phecode 1.2 mapping tables. It is well established that there is no association between HIV status and colorectal cancer.[47,48] We investigated the underlying ICD codes that are qualify as a colorectal cancer case. Our analyses in the manuscript use the phecode mapping table present in the PheWAS R package (version 1.2).[49,50] We also show qualifying ICD codes for a different phecode mapping table (version X),[51,52] which defines over 3,600 traits. The results of the differences in qualifying ICD codes, number of individuals with the ICD code, and the number (and percent) overlap with individuals who have HIV according to their version 1.2 defined phecode are summarized in **Table S5**. We see that there is significant overlap between individuals with ICD codes for anal Pap smears, inconclusive results and *carcinoma in situ* and HIV status. These codes are present in the version 1.2 mapping table, but not in the version X mapping table. Codes present in the version 1.2 definition also include malignant neoplasms of the anus, but not in the version X definition. And there is evidence that people living with HIV experience higher incidence of anal cancer.[53] Because version X has more traits, there is greater separation between colorectal cancer and anal cancer.

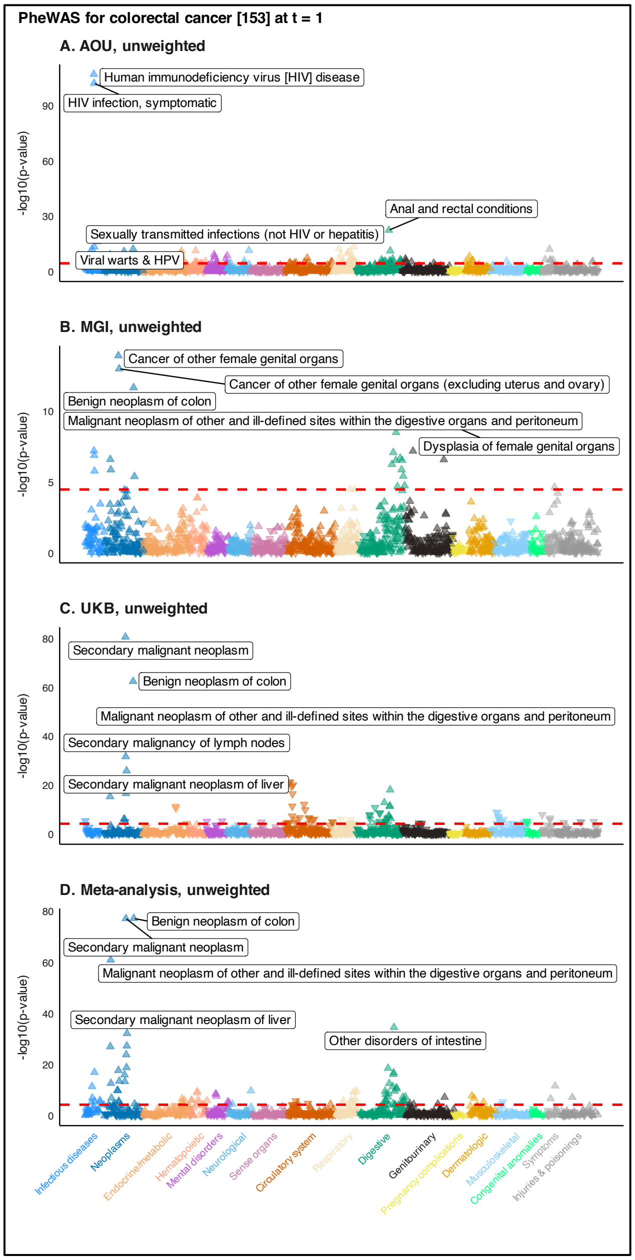


**Figure S14**. Manhattan plots summarizing unweighted (panels A-C) phenomewide association studies for colorectal cancer in All of Us, the Michigan Genomics Initiative, and UK Biobank using 1:2 case:non-case matched data restricted to one year prior to initial diagnosis. Panel D shows the unweighted meta-analysis PheWAS, respectively. The dashed red line represents the Bonferroni-corrected p-value threshold (-log10(0.05/number of traits)). The five traits with the smallest p-values are labeled. The upward (downward) orientation of the triangle indicates a positive (negative) association.

**Table S5**. Comparison between ICD codes by colorectal cancer phecode mapping table, count with ICD code, and overlap with individuals who have HIV phecode (sorted by proportion of overlap).

| | | | Qualifying ICD codes for colorectal cancer phecode by mapping table version | | | | |
|---|---|---|---|---|---|---|---|
| **ICD Code** | **ICD Version** | **ICD Code Description** | **Version 1.2** | **Version X** | **Number with qualifying ICD code** | **Number with ICD codes and HIV phecode** | **Percent** |
| 796.71 | ICD9CM | Papanicolaou smear of anus with atypical squamous cells of undetermined significance (ASC-US) | 1 | 0 | 87 | 74 | 85.1 |
| 796.74 | ICD9CM | Papanicolaou smear of anus with high grade squamous intraepithelial lesion (HGSIL) | 1 | 0 | 29 | 23 | 79.3 |
| 796.73 | ICD9CM | Papanicolaou smear of anus with low grade squamous intraepithelial lesion (LGSIL) | 1 | 0 | 100 | 79 | 79 |
| 230.6 | ICD9CM | Carcinoma in situ of anus, unspecified | 1 | 0 | 70 | 53 | 75.7 |
| R85.613 | ICD10CM | High grade squamous intraepithelial lesion on cytologic smear of anus (HGSIL) | 1 | 0 | 61 | 45 | 73.8 |
| R85.612 | ICD10CM | Low grade squamous intraepithelial lesion on cytologic smear of anus (LGSIL) | 1 | 0 | 166 | 118 | 71.1 |
| 796.7 | ICD9CM | Abnormal glandular Papanicolaou smear of anus | 1 | 0 | 124 | 86 | 69.4 |
| R85.610 | ICD10CM | Atypical squamous cells of undetermined significance on cytologic smear of anus (ASC-US) | 1 | 0 | 155 | 105 | 67.7 |
| 230.5 | ICD9CM | Carcinoma in situ of anal canal | 1 | 0 | 74 | 50 | 67.6 |
| R85.619 | ICD10CM | Unspecified abnormal cytological findings in specimens from anus | 1 | 0 | 103 | 61 | 59.2 |
| D01.3 | ICD10CM | Carcinoma in situ of anus and anal canal | 1 | 0 | 219 | 129 | 58.9 |
| 796.72 | ICD9CM | Papanicolaou smear of anus with atypical squamous cells cannot exclude high grade squamous intraepithelial lesion (ASC-H) | 1 | 0 | 15 | 8 | 53.3 |
| C7A.024 | ICD10CM | Malignant carcinoid tumor of the descending colon | 1 | 0 | 2 | 1 | 50 |
| R85.611 | ICD10CM | Atypical squamous cells cannot exclude high grade squamous intraepithelial lesion on cytologic smear of anus (ASC-H) | 1 | 0 | 29 | 14 | 48.3 |
| 796.76 | ICD9CM | Papanicolaou smear of anus with cytologic evidence of malignancy | 1 | 0 | 3 | 1 | 33.3 |
| 154.3 | ICD9CM | Malignant neoplasm of anus, unspecified site | 1 | 0 | 107 | 20 | 18.7 |
| C21.0 | ICD10CM | Malignant neoplasm of anus, unspecified | 1 | 0 | 163 | 28 | 17.2 |
| 209.1 | ICD9CM | Malignant carcinoid tumor of the large intestine, unspecified portion | 1 | 0 | 7 | 1 | 14.3 |
| C21.1 | ICD10CM | Malignant neoplasm of anal canal | 1 | 0 | 96 | 13 | 13.5 |
| D37.5 | ICD10CM | Neoplasm of uncertain behavior of rectum | 0 | 1 | 16 | 2 | 12.5 |
| 154.2 | ICD9CM | Malignant neoplasm of anal canal | 1 | 0 | 56 | 6 | 10.7 |
| 154.8 | ICD9CM | Malignant neoplasm of other sites of rectum, rectosigmoid junction, and anus | 1 | 0 | 30 | 2 | 6.7 |
| C21.8 | ICD10CM | Malignant neoplasm of overlapping sites of rectum, anus and anal canal | 1 | 0 | 45 | 3 | 6.7 |
| 153.4 | ICD9CM | Malignant neoplasm of cecum | 1 | 1 | 71 | 4 | 5.6 |
| 154.1 | ICD9CM | Malignant neoplasm of rectum | 1 | 1 | 204 | 11 | 5.4 |
| C18.8 | ICD10CM | Malignant neoplasm of overlapping sites of colon | 1 | 1 | 45 | 2 | 4.4 |
| C18.4 | ICD10CM | Malignant neoplasm of transverse colon | 1 | 1 | 69 | 3 | 4.3 |
| C20 | ICD10CM | Malignant neoplasm of rectum | 1 | 1 | 362 | 15 | 4.1 |
| C18.5 | ICD10CM | Malignant neoplasm of splenic flexure | 1 | 1 | 30 | 1 | 3.3 |
| 153.6 | ICD9CM | Malignant neoplasm of ascending colon | 1 | 1 | 125 | 4 | 3.2 |
| 153.1 | ICD9CM | Malignant neoplasm of transverse colon | 1 | 1 | 35 | 1 | 2.9 |
| D37.3 | ICD10CM | Neoplasm of uncertain behavior of appendix | 0 | 1 | 37 | 1 | 2.7 |
| 153.5 | ICD9CM | Malignant neoplasm of appendix vermiformis | 1 | 1 | 42 | 1 | 2.4 |
| C19 | ICD10CM | Malignant neoplasm of rectosigmoid junction | 1 | 1 | 302 | 7 | 2.3 |
| C18.3 | ICD10CM | Malignant neoplasm of hepatic flexure | 1 | 1 | 47 | 1 | 2.1 |
| D37.4 | ICD10CM | Neoplasm of uncertain behavior of colon | 0 | 1 | 48 | 1 | 2.1 |
| C18.9 | ICD10CM | Malignant neoplasm of colon, unspecified | 1 | 1 | 979 | 18 | 1.8 |
| C18.2 | ICD10CM | Malignant neoplasm of ascending colon | 1 | 1 | 238 | 4 | 1.7 |
| 153.3 | ICD9CM | Malignant neoplasm of sigmoid colon | 1 | 1 | 125 | 2 | 1.6 |
| 153.9 | ICD9CM | Malignant neoplasm of colon, unspecified site | 1 | 1 | 632 | 10 | 1.6 |
| 153.8 | ICD9CM | Malignant neoplasm of other specified sites of large intestine | 1 | 1 | 69 | 1 | 1.4 |
| 154 | ICD9CM | Malignant neoplasm of rectosigmoid junction | 1 | 1 | 167 | 2 | 1.2 |
| C18 | ICD10CM | Malignant neoplasm of colon | 1 | 1 | 484 | 6 | 1.2 |
| C18.1 | ICD10CM | Malignant neoplasm of appendix | 1 | 1 | 89 | 1 | 1.1 |
| C18.7 | ICD10CM | Malignant neoplasm of sigmoid colon | 1 | 1 | 231 | 1 | 0.4 |
| 153 | ICD9CM | Malignant neoplasm of hepatic flexure | 1 | 1 | 40 | 0 | 0 |
| 153.2 | ICD9CM | Malignant neoplasm of descending colon | 1 | 1 | 37 | 0 | 0 |
| 153.7 | ICD9CM | Malignant neoplasm of splenic flexure | 1 | 1 | 13 | 0 | 0 |
| 154 | ICD9CM | Malignant neoplasm of rectum rectosigmoid junction and anus | 1 | 0 | 6 | 0 | 0 |
| 159 | ICD9CM | Malignant neoplasm of intestinal tract, part unspecified | 1 | 0 | 38 | 0 | 0 |
| 209.1 | ICD9CM | Malignant carcinoid tumors of the appendix, large intestine, and rectum | 1 | 0 | 0 | 0 | 0 |
| 209.11 | ICD9CM | Malignant carcinoid tumor of the appendix | 1 | 0 | 8 | 0 | 0 |
| 209.12 | ICD9CM | Malignant carcinoid tumor of the cecum | 1 | 0 | 5 | 0 | 0 |
| 209.13 | ICD9CM | Malignant carcinoid tumor of the ascending colon | 1 | 0 | 0 | 0 | 0 |
| 209.14 | ICD9CM | Malignant carcinoid tumor of the transverse colon | 1 | 0 | 0 | 0 | 0 |
| 209.15 | ICD9CM | Malignant carcinoid tumor of the descending colon | 1 | 0 | 1 | 0 | 0 |
| 209.16 | ICD9CM | Malignant carcinoid tumor of the sigmoid colon | 1 | 0 | 1 | 0 | 0 |
| 209.17 | ICD9CM | Malignant carcinoid tumor of the rectum | 1 | 0 | 5 | 0 | 0 |
| 230.3 | ICD9CM | Carcinoma in situ of colon | 1 | 1 | 71 | 0 | 0 |
| 230.4 | ICD9CM | Carcinoma in situ of rectum | 1 | 1 | 6 | 0 | 0 |
| 796.7 | ICD9CM | Abnormal cytologic smear of anus and anal HPV | 1 | 0 | 1 | 0 | 0 |
| C18.0 | ICD10CM | Malignant neoplasm of cecum | 1 | 1 | 115 | 0 | 0 |
| C18.6 | ICD10CM | Malignant neoplasm of descending colon | 1 | 1 | 79 | 0 | 0 |
| C21 | ICD10CM | Malignant neoplasm of anus and anal canal | 1 | 0 | 0 | 0 | 0 |
| C21.2 | ICD10CM | Malignant neoplasm of cloacogenic zone | 1 | 0 | 2 | 0 | 0 |
| C26.0 | ICD10CM | Malignant neoplasm of intestinal tract, part unspecified | 1 | 0 | 16 | 0 | 0 |
| C7A.020 | ICD10CM | Malignant carcinoid tumor of the appendix | 1 | 0 | 21 | 0 | 0 |
| C7A.021 | ICD10CM | Malignant carcinoid tumor of the cecum | 1 | 0 | 6 | 0 | 0 |
| C7A.022 | ICD10CM | Malignant carcinoid tumor of the ascending colon | 1 | 0 | 1 | 0 | 0 |
| C7A.023 | ICD10CM | Malignant carcinoid tumor of the transverse colon | 1 | 0 | 1 | 0 | 0 |
| C7A.025 | ICD10CM | Malignant carcinoid tumor of the sigmoid colon | 1 | 0 | 2 | 0 | 0 |
| C7A.026 | ICD10CM | Malignant carcinoid tumor of the rectum | 1 | 0 | 16 | 0 | 0 |
| C7A.029 | ICD10CM | Malignant carcinoid tumor of the large intestine, unspecified portion | 1 | 0 | 8 | 0 | 0 |
| D01.0 | ICD10CM | Carcinoma in situ of colon | 1 | 1 | 38 | 0 | 0 |
| D01.1 | ICD10CM | Carcinoma in situ of rectosigmoid junction | 1 | 1 | 0 | 0 | 0 |
| D01.2 | ICD10CM | Carcinoma in situ of rectum | 1 | 1 | 6 | 0 | 0 |
| R85.614 | ICD10CM | Cytologic evidence of malignancy on smear of anus | 1 | 0 | 0 | 0 | 0 |
| V10.05 | ICD9CM | Personal history of malignant neoplasm of large intestine | 1 | 1 | 0 | 0 | 0 |
| V10.06 | ICD9CM | Personal history of malignant neoplasm of rectum, rectosigmoid junction, and anus | 1 | 0 | 0 | 0 | 0 |
| Z85.02 | ICD10CM | Personal history of malignant neoplasm of stomach | 1 | 0 | 0 | 0 | 0 |
| Z85.03 | ICD10CM | Personal history of malignant neoplasm of large intestine | 1 | 1 | 0 | 0 | 0 |
| Z85.038 | ICD10CM | Personal history of other malignant neoplasm of large intestine | 1 | 1 | 0 | 0 | 0 |
| Z85.048 | ICD10CM | Personal history of other malignant neoplasm of rectum, rectosigmoid junction, and anus | 1 | 0 | 0 | 0 | 0 |

We also present the 2x2 tables and crude odds ratios between these two different definitions of colorectal cancer and HIV status in **Table S6**. The crude odds ratio in the time-restricted phenome (t = 1) is 10.29 using version 1.2 colorectal cancer mapping and 0.32 using version X mapping.

**Table S6**. Colorectal cancer and HIV contingency table by phecode mapping version and crude odds ratio

| | **Colorectal cancer** | | | |
|---|---|---|---|---|
| | **Version 1.2** | | **Version X** | |
| **HIV** | 1 | 0 | 1 | 0 |
| 1 | 478 | 108 | 60 | 526 |
| 0 | 2611 | 6069 | 2305 | 6375 |
| **Crude odds ratio** | 10.29 | | 0.32 | |

Other hits in the infectious disease category like HIV infection, symptomatic (phecode 071.1) and viral warts & HPV (phecode 078) share many of the same underlying ICD codes as with phecode 071, which implies similar overlap with colorectal cancer as defined by phecode mapping table version 1.2.

12. White A, Ironmonger L, Steele RJC, Ormiston-Smith N, Crawford C, Seims A. A review of sex-related differences in colorectal cancer incidence, screening uptake,

routes to diagnosis, cancer stage and survival in the UK. *BMC Cancer*. 2018;18(1):906. doi:10.1186/s12885-018-4786-7
